# Tool-wielding language-model-based agent offers conversational exploration of clinical tabular data

**DOI:** 10.64898/2025.12.01.25341392

**Authors:** Andrew Yang, Joshua Woo, Ryan Zhang, Alan Mach, Prem Ramkumar, Ying Ma

**Affiliations:** The Warren Alpert Medical School of Brown University, Providence, RI 02903, USA; Department of Statistics and Data Sciences, University of Texas at Austin, Austin, TX 78712, USA; Commons Clinic, Long Beach, CA 90808, USA; Department of Biostatistics, Brown University, Providence, RI 02903, USA; Center for Computational Molecular Biology, Brown University, Providence, RI 02903, USA

## Abstract

Advancing evidence-based medicine requires integrating clinical expertise with data analysis. While clinicians contribute essential domain knowledge, applying modern data science methods often requires specialized training, creating a barrier to adoption. To bridge this gap, we developed ChatDA, an artificial intelligence agent enabling large-language-model-mediated conversational analysis of de-identified clinical tabular datasets. ChatDA empowers clinicians to extract meaningful insights efficiently and accurately, making data-driven clinical research more accessible and effective.

## Main

Data analysis is fundamental to evidence-based medicine, supporting both clinical decision-making and the interpretation of research findings^1–3^. However, many clinicians may face technical challenges when analyzing data, as their training often emphasizes clinical practice over statistics and data science^4–6^. These challenges may lead to inefficiencies or reliance on external data analysts, preventing clinical experts from fully engaging with their data and potentially slowing the translation of data-driven insights into actionable interventions. Recent advancements in artificial intelligence (AI), particularly large language models (LLMs), offer a promising path toward lowering these barriers by expanding access to data science capabilities among clinicians.

LLMs have demonstrated remarkable potential across various healthcare applications, from bioinformatics research assistance^7,8^ to AI-assisted clinical decision-making^9,10^. These models excel in domain knowledge recall^11^, information retrieval^12,13^, logical reasoning^14^, and code generation^15^. Particularly relevant to clinical research assistance, AI-powered code-writing agents have proven highly effective for data analysis tasks from statistical testing to machine learning^7,16–18^. While such agents hold promise for AI-assisted clinical data analysis, their effectiveness hinges on the capabilities of powerful LLMs, as smaller models struggle with accurately generating complex code scripts^17,18^. These high-performance LLMs rely on cloud-based inference, raising data privacy concerns: when code-writing agents process clinical data, they may inadvertently expose sensitive patient information to external providers^19^. Even if the data is de-identified, the sharing of detailed patient-level information increases the risk of re-identification^20,21^, posing significant ethical and regulatory challenges. This underscores the tension between leveraging AI in clinical research and maintaining data protection standards.

To address these challenges, we developed Chat Data Analyst (ChatDA), an AI agent designed for analyzing de-identified tabular clinical data that operates through specialized tool use rather than code generation. ChatDA reduces the data privacy risks associated with conventional code-writing agents by restricting the language model’s access to data: analyses are conducted through custom tools that return only population-level insights, ensuring individual-level data remains inaccessible to the underlying cloud-hosted LLM. Encouraged by recent studies demonstrating the effectiveness of agentic tool use in practical applications^22–27^, we hypothesized that ChatDA, similarly equipped with a specialized toolkit, could match or outperform state-of-the-art code-execution agents. Moreover, this approach may offer additional advantages: by guiding analyses through well-defined, pre-validated operations instead of free-form code execution, it enhances result consistency, reduces the risk of errors, and improves interpretability.

In this study, we evaluate our hypothesis by benchmarking ChatDA across a range of standard clinical data analysis tasks, including summary statistics generation, statistical testing, regression analysis, and machine learning modeling. We compare its performance to four existing agents on 21 publicly available datasets: (1) OpenAI Advanced Data Analysis (ADA), a ChatGPT feature that enables OpenAI models to execute code to analyze user-uploaded data; (2) Open Interpreter, an open-source coding interface for LLMs; (3) Data Interpreter^17^, an open-source agent that tackles complex data analysis tasks primarily through code generation; and (4) LAMBDA^28^, a peer-reviewed coding agent designed for data analysis. Then, to demonstrate the practical utility of ChatDA, we present a case study in which ChatDA analyzes a proprietary, de-identified hip arthroplasty dataset to extract meaningful population-level insights.

ChatDA, illustrated in Figure 1A, is an LLM-powered agent that operates a custom data science toolkit. ChatDA’s toolkit—an extension of TableMage, a novel software package for low-code clinical data science—provides an integrated data and analysis environment, supporting data transformation, statistical testing, figure generation, regression analysis, and machine learning modeling (Methods). ChatDA can optionally operate a Python interpreter, which can be enabled to boost local LLM-powered agent performance. When powered by a multimodal large language model such as GPT-4o, ChatDA can analyze the figures it generates, further enhancing its capability to interpret results.

**Figure 1.**
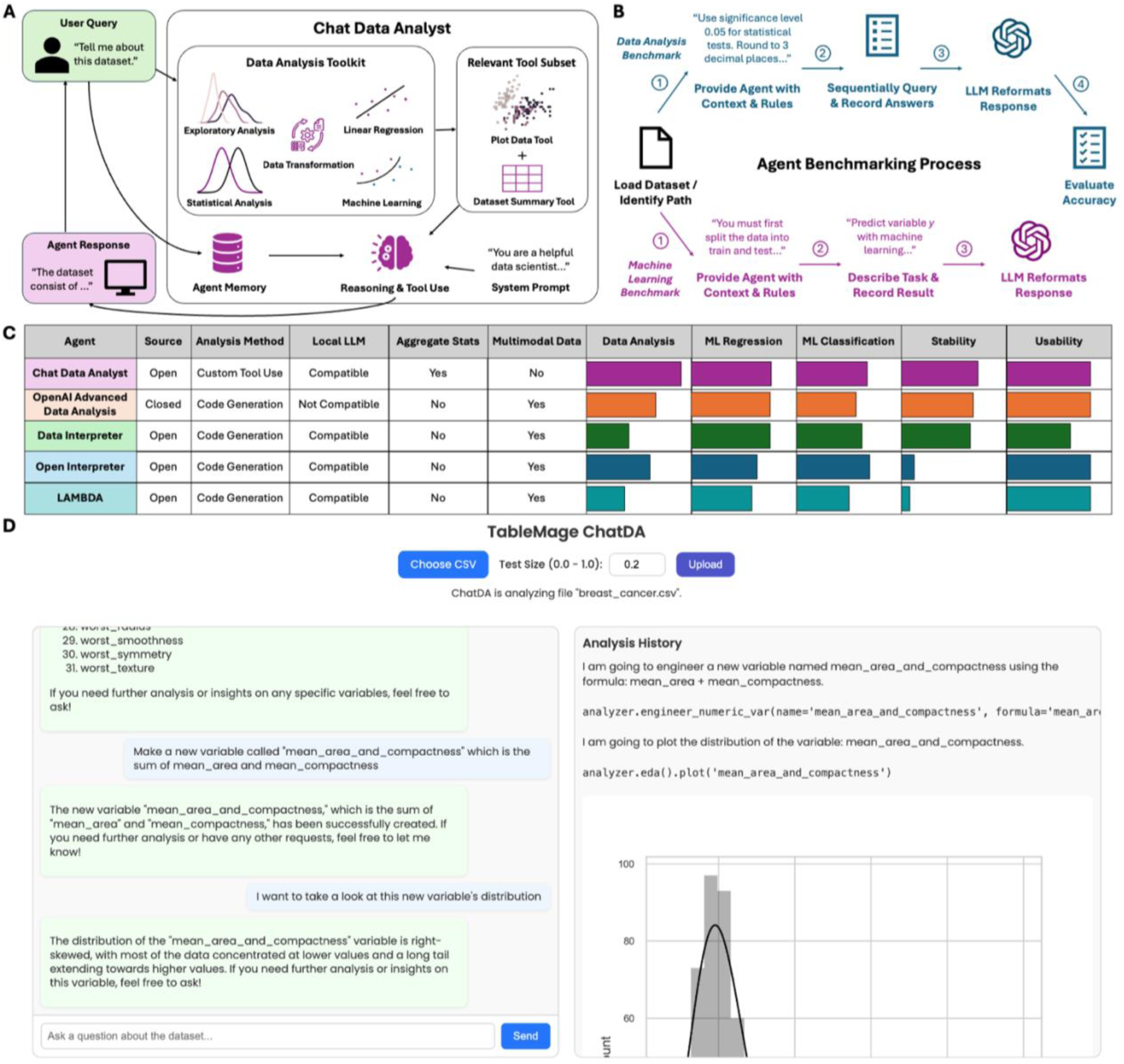
Chat Data Analyst overview and evaluation. **(A)** Chat Data Analyst (ChatDA) is a large-language-model-powered AI agent specialized in clinical data science tasks, including exploratory data analysis, statistical testing, regression analysis, and machine learning modeling. ChatDA operates via a custom toolkit built on TableMage, a novel, low-code clinical data analysis software package. **(B)** Schematic of the agent benchmarking process for evaluating data analysis accuracy and machine learning performance. **(C)** Summary comparison of four AI-based data analysis agents: ChatDA (purple), OpenAI Advanced Data Analysis (orange), Data Interpreter (green), Open Interpreter (blue), and LAMBDA (turquoise). Detailed benchmarking results are provided in Supplementary Figures 1-3. **(D)** Example of ChatDA performing conversational data analysis on a publicly available breast cancer classification dataset. The ChatDA user interface displays the user’s conversation on the left and ChatDA’s analytical workflow on the right. Corresponding TableMage code segments ensure reproducibility of tool-based actions.

To evaluate ChatDA’s accuracy in common data analysis tasks, we curated a novel benchmark for assessing AI agents in data workflows (Methods, Figure 1B). We compared ChatDA against OpenAI ADA, Open Interpreter, Data Interpreter, and LAMBDA using this benchmark. ChatDA achieved the highest accuracy and stability across all question topics with a 10% to 18% improvement in accuracy over the next-best agent. All reported accuracy metrics are accompanied by 95% confidence intervals to quantify performance variability, and standard errors were computed across repeated trials to account for response fluctuation inherent in stochastic LLM behavior. Specifically, ChatDA achieved an overall accuracy of 0.951 (95% CI: ± 0.008), compared to 0.830 ± 0.010 for OpenAI ADA, 0.803 ± 0.029 for Open Interpreter, 0.701 ± 0.011 for Data Interpreter, and 0.599 ± 0.035 for LAMBDA (Figure 1C, Supplementary Figure 1). Notably, ChatDA exhibited greater output stability than other agents, as reflected in its narrower confidence intervals across evaluation runs. This robustness addresses a well-documented challenge in LLM-powered agents—namely, the variability and unpredictability in generated outputs under identical prompts. By relying on structured tool-based workflows rather than unconstrained code generation, ChatDA reduces variance in performance and delivers more consistent, interpretable results. ChatDA also outperformed other code-writing agents in retaining data transformations, such as scaled or engineered features, between analysis steps (ChatDA: 0.887 ± 0.024; OpenAI ADA: 0.692 ± 0.030; Open Interpreter: 0.696 ± 0.050; Data Interpreter: 0.296 ± 0.019; LAMBDA: 0.479 ± 0.069; see Supplementary Figure 1), likely due to its data-integrated toolkit, which automatically preserves transformations for future tasks.

Next, we evaluated ChatDA’s capability to train machine learning models across a set of publicly curated regression and classification tasks for tabular datasets (Methods). ChatDA achieved competitive performance on the machine learning benchmark, outperforming OpenAI ADA in 10 out of 11 tasks (i.e., achieving a lower average test RMSE for regression or a higher average test AUC for classification), Open Interpreter in 6 out of 11 tasks, Data Interpreter in 9 out of 11 tasks, and LAMBDA in 7 out of 11 tasks. ChatDA’s performance on both the data analysis benchmark and the machine learning benchmark demonstrates its potential to match or surpass existing state-of-the-art conversational data analysis solutions at machine learning modeling. Complete machine learning benchmark results are presented in Supplementary Figures 2-3.

To demonstrate ChatDA in practice, we conducted a conversational case study using real-world de-identified clinical data (Methods). Through the ChatDA user interface (Figure 1D), we analyzed an in-house proprietary dataset of 1,419 knee arthroplasty patients to examine whether quantitative anatomical and functional features^29^ could predict the surgeon-selected procedure—Total Knee Arthroplasty (TKA) or Unicompartmental Knee Arthroplasty (UKA). ChatDA first summarized the dataset, removed cases with missing knee arthroplasty annotations, and performed imputation using median values for numeric features and a “missing” category for categorical ones. Among 35 potential predictors, ChatDA applied the Boruta method^30^ and identified 6 top features: joint line convergence angle, lateral tibial width, medial proximal tibial angle, tibiofemoral angle, tibial width, and tibial tubercle-trochlear groove distance. A logistic regression model trained on these features achieved a test area under the receiver operating characteristic (AUC) of 0.632, suggesting moderate predictive power. Joint line convergence angle, tibiofemoral angle, and tibial tubercle-trochlear groove distance emerged as statistically significant predictors. Notably, ChatDA found that for every unit increase in joint line convergence angle, the odds of UKA increased by 14.3%. Although random forest and XGBoost models were also evaluated, they did not outperform logistic regression but reaffirmed the importance of tibial tubercle-trochlear groove distance and tibiofemoral angle. All results were generated in a single uninterrupted conversational session without manual correction, highlighting ChatDA’s usability and robustness in a clinical research context Figure 2B-2D. The complete conversation transcript is available in Supplementary Note 6.

**Figure 2.**
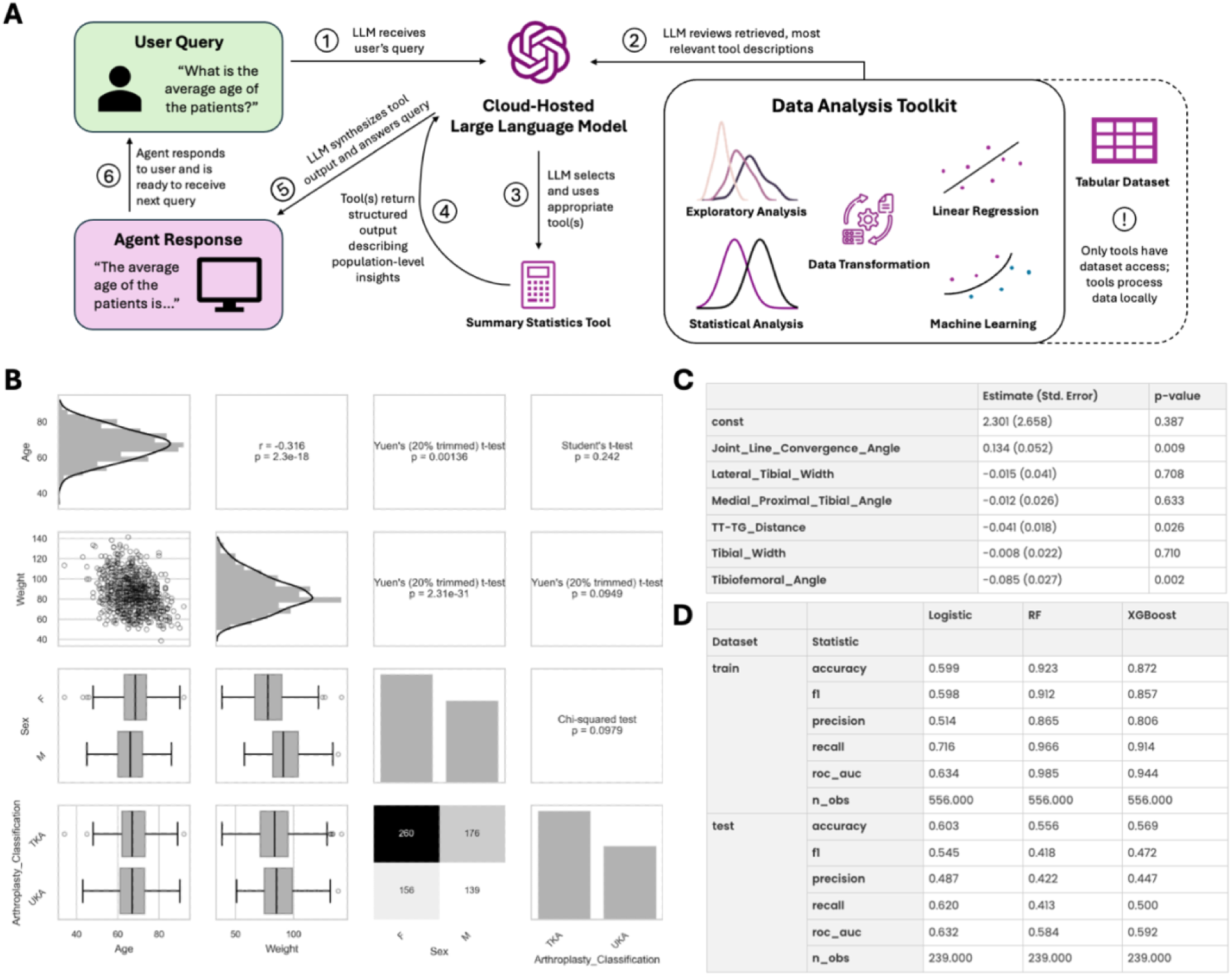
Case study insights: analysis of a real-world, proprietary knee arthroplasty preoperative decision-making dataset. **(A)** Schematic showcasing ChatDA’s tools-only data analysis mode. The schematic illustrates that ChatDA’s underlying LLM cannot directly access individual-level data. ChatDA, set to tools-only mode, was used to analyze a de-identified knee arthroplasty dataset. **(B)** ChatDA-generated figure depicting pairwise relationships between age, weight, sex, and knee arthroplasty type. Box plots display medians (center line), quartiles (box limits), and 1.5× IQR whiskers; outliers are shown as individual points. **(C)** ChatDA-generated table of logistic regression coefficients. **(D)** ChatDA-generated table of machine learning model performances.

ChatDA addresses a critical challenge in AI-assisted de-identified clinical data analysis: enhancing patient data privacy. Traditional de-identification techniques are often insufficient when the individual-level data is passed to cloud-hosted LLMs^21,31^: even after explicit identifiers are removed, individual-level data points can still lead to re-identification risks^32^. ChatDA introduces two mechanisms to reduce the risk of re-identification.

1. **Local LLM deployment.** ChatDA is compatible with locally hosted open-source LLMs, such as Llama models^33,34^, allowing users to perform AI-assisted data analysis entirely on their own hardware. This approach ensures that no patient data leaves the local environment, completely eliminating external exposure risks. However, running high-performance LLMs locally requires substantial computational resources, making this option inaccessible to many due to hardware constraints.
2. **Restrictive tools-only analysis mode.** To accommodate users reliant on cloud-hosted LLMs, ChatDA provides an alternative layer of re-identification risk reduction. As demonstrated in both the benchmarking study (Supplementary Figures 1-3) and the case study (Figure 2), ChatDA retains most of its functionality even when its Python interpreter is disabled. Granting a cloud-based LLM direct access to a coding environment containing de-identified clinical data poses a significant privacy risk, as the model could generate code that inadvertently exposes individual-level records, increasing the risk of re-identification. By removing Python interpreter access, ChatDA is explicitly prevented from indexing into the dataset or accessing raw patient data. Instead, all computations are performed locally, and only aggregated, population-level results—such as variable names, summary statistics, statistical test results, and model performance metrics—are passed to the LLM (Figure 2A). When exclusively analyzing de-identified datasets in tools-only mode, ChatDA tool outputs align with standard reporting practices in clinical research and reduce the likelihood of compromising patient confidentiality^35,36^ when compared to code-generating agents.

This work has several limitations that warrant discussion. First, while ChatDA’s “tools-only” mode reduces re-identification risk by preventing cloud LLMs from accessing individual-level records, this approach does not eliminate all privacy concerns; modern privacy research has shown that aggregate statistics can still be vulnerable to re-identification^37–41^. Therefore, ChatDA is intended for use only with datasets that have been properly de-identified in accordance with applicable legal standards (see Supplementary Note 1 for additional discussion and practical guidance). Second, the scope of ChatDA is currently limited to tabular datasets, a focus that does not address the challenges of analyzing other common clinical data types like medical imaging or unstructured text; moreover, without its Python interpreter, ChatDA’s functionality is strictly limited to the availability of pre-defined tools (Supplementary Note 2). Third, our accuracy evaluation was performed on a custom-designed DataAnalysisQA benchmark, which may introduce inherent biases; comprehensively benchmarking data analysis agents remains an open challenge (Supplementary Note 3), and additional analyses such as error propagation could further enhance agent evaluation. Fourth, the practical utility of ChatDA is demonstrated through only a single case study, and a formal user study with multiple clinicians is needed to fully validate its real-world effectiveness and usability. Finally, this study compares ChatDA only with code-writing agents and human-generated Python code; as new domain-specific low-code and no-code tools emerge for clinical data analysis, future evaluations should also include those systems.

By reducing barriers to data analysis and enabling secure AI-mediated workflows, ChatDA empowers healthcare professionals to extract meaningful insights from de-identified tables and advance evidence-based medicine. Data analysis agents like ChatDA have the potential to transform quantitative clinical research by accelerating discovery, broadening access to data-driven insights, and strengthening the foundation for clinical decision-making in an increasingly data-rich environment. ChatDA’s consistent performance across both benchmark evaluations and real-world applications further establishes it as a reliable and trustworthy tool for data analysis.

## Methods

### Agent development

To enable AI-assisted clinical data analysis primarily through predefined tools, we developed TableMage, a high-level, low-code Python package designed specifically for analyzing tabular clinical data. Our focus on tabular formats reflects their central role in clinical research (Supplementary Note 4). TableMage integrates functionality from widely used packages in the Python ecosystem including NumPy^42^ and pandas^43^ for data manipulation, SciPy^44^ and Statsmodels^45^ for statistical analysis, Matplotlib^46^ and seaborn^47^ for data visualization, Scikit-learn^48^ and Optuna^49^ for machine learning and hyperparameter optimization, and Boruta^30^ for feature selection.

A core feature of TableMage is its seamless integration of data exploration, transformation, and modeling within a unified framework. It enforces strict separation between training and testing sets by requiring a train-test split prior to any analysis. This guarantees that the test set remains entirely isolated from all transformation and modeling processes, effectively mitigating data leakage and preserving the integrity of model assessment. This design allows users—whether human or AI agent—to freely conduct exploratory data analysis, apply transformations, and train models without worrying about execution order or inadvertently neglecting to persist transformations. By automatically tracking and applying transformations, TableMage ensures consistency across training and testing data, preventing common errors such as applying scaling only to the training set or failing to carry over preprocessing steps to new data. We note that exploratory analyses—summary statistics generation, distribution visualization, statistical testing, and correlation analyses—are still performed on the entire dataset, and the initial train-test split is enforced primarily to ensure proper modeling, as preprocessing-related mistakes are frequent (https://scikit-learn.org/stable/common_pitfalls.html^48^). Additionally, TableMage enables highly efficient pipelining by automatically converting any sequence of transformations into a custom scikit-learn pipeline. When combined with a fitted model, this pipeline can be serialized for downstream analysis and deployment, streamlining the entire machine learning workflow.

Building upon TableMage, we developed a suite of clinical data analysis tools using LlamaIndex^50^, an AI agent development framework. These tools were then integrated into Chat Data Analyst (ChatDA), a function-calling agent designed to facilitate conversational data analysis. To optimize the use of the underlying LLM’s context window, ChatDA dynamically selects only the most relevant tools for a given query through retrieval-augmented generation^13^. Once the appropriate tools are identified, ChatDA executes them and synthesizes a response based on their outputs. If appropriate, ChatDA can use multiple tools to respond to a single query. As a conversational agent, ChatDA utilizes a buffer-based memory for short-term recall and a vector index-based memory for long-term retention, ensuring continuity and context awareness in conversations.

ChatDA optionally provides access to a Python interpreter for short-term tasks such as data indexing and generating custom plots. However, because the Python interpreter allows direct access to individual-level data, its use must be restricted to non-sensitive datasets or cases where the underlying LLM is locally hosted to ensure data security. Even without access to its Python interpreter, ChatDA can still perform operations such as categorical variable encoding, value counting, and min/max value reporting using its built-in tools, thereby limiting its applicability to properly de-identified datasets. We provide a discussion of the aggregate-level reporting scheme of ChatDA’s tools and its potential to reduce the likelihood of re-identification in Supplementary Note 1.

ChatDA is compatible with cloud-hosted LLMs (closed-source or open-source) as well as with locally hosted open-source LLMs. If ChatDA is powered by a multimodal foundation language model such as GPT-4o, it can view and analyze the figures generated by its tools. ChatDA’s tools are listed in Supplementary Table 1.

### Agent benchmarking

We evaluated the performance of ChatDA, OpenAI Advanced Data Analysis, Open Interpreter, and Data Interpreter on a range of non-machine learning (non-ML) and supervised machine learning (ML) tasks.

### Non-ML Benchmark: DataAnalysisQA

To assess text in, text out artificial intelligence systems (e.g., language-model-based data analysis/coding agents) on foundational data analysis capabilities, we developed DataAnalysisQA, a benchmark consisting of 100 question-answer pairs across 10 publicly available tabular datasets. This benchmark was inspired by InfiAgent-DABench^18^ but addresses several key limitations we observed in its design:

1. **Code-specific technical language.** Many questions in InfiAgent-DABench prompt agents to invoke specific Python functions, favoring Python code-writing agents rather than general-purpose data analysts.
2. **Dependence on OpenAI Advanced Data Analysis.** InfiAgent-DABench was constructed using GPT-4 and OpenAI ADA, making it less suitable for independently evaluating OpenAI’s own tools.
3. **Lack of continuity.** All questions in InfiAgent-DABench are independent, limiting its ability to assess whether agents can retain and apply transformations across sequential queries.

To address these limitations, we designed DataAnalysisQA using human-generated questions and ground truth answers, ensuring broader applicability and independence from specific AI systems. For each of the 10 datasets, we curated a coherent sequence of queries covering summary statistics, data transformation, indexing, statistical testing, feature engineering, and regression. This sequential structure enabled the evaluation of each agent’s ability to retain and apply data transformations over multiple interactions. Unlike the code-specific instructions commonly found in InfiAgent-DABench, our benchmark queries were designed to be accessible while maintaining clarity and ensuring unique, well-defined answers. Ground truth results were generated manually for each query, and all question-answer pairs were labeled with one or more topic categories, including indexing, statistical testing, data transformation, persistence (retention of previous transformations), summary statistics, and regression analysis. In total, DataAnalysisQA consists of 100 questions across these categories.

### Machine Learning Benchmark

In addition to non-ML tasks, we evaluated all agents on 11 publicly available datasets from a tabular ML benchmark^51^ on OpenML^52^. The 11 datasets represent the subset of the tabular data machine learning benchmark with no more than 10,000 rows and no more than 100 columns; larger datasets were filtered out to ensure fair evaluation of OpenAI Advanced Data Analysis, which is the limited by its cloud CPU memory and runtime capacities. Each agent was instructed or preconfigured to perform a 70-30 train-test split with identical random seeds and to report the best-performing model based on evaluation metrics. For regression tasks, we used root mean squared error (RMSE) as the primary metric, while for binary classification tasks, we evaluated model performance using the area under the receiver operating characteristic curve (AUC).

For both ML and non-ML benchmarks, ChatDA was evaluated in two modes: (1) normal mode, where it had access to both its custom tools and a Python interpreter, and (2) tools-only mode, where it was restricted to its custom tools only. In normal mode, ChatDA was tested with both a closed-source LLM (GPT-4o) and an open-source LLM (Llama 3.3 70B). While normal mode ChatDA does not reduce data re-identification risk when using a cloud hosted LLM, it does guarantee data privacy when powered by a locally hosted LLM. Notably, Llama 3.3 70B can be deployed on consumer-grade hardware, offering a privacy-preserving alternative. In tools-only mode, ChatDA was tested exclusively with the closed-source GPT-4o. Together, these evaluations assessed two approaches to reducing the risk of data re-identification: (1) local LLM usage (normal mode with Llama 3.3 70B) and (2) a non-coding approach (tools-only mode with GPT-4o). Additionally, normal mode with GPT-4o was tested as an option for non-sensitive datasets.

For both benchmarks, agents return unstructured output. Following InfiAgent-DABench, we employed LLMs to extract structured answers from unstructured output. To assess stability, each agent was tested across 10 replicates in both the data analysis and machine learning benchmarks. For each metric, the mean and its 95% confidence interval were reported. Complete DataAnalysisQA and machine learning benchmarking results are presented in Supplementary Figures 1-3. For DataAnalysisQA, failure rates, defined as the proportion of questions an agent could not answer, were calculated by category for each agent (Supplementary Figure 4). For ChatDA, tool-level error rates were determined by manually labeling each question with the tool expected to produce the correct answer (Supplementary Figure 5). We include ChatDA’s performance on the InfiAgent-DABench benchmark in Supplementary Note 5. In addition to the performance benchmarks, a brief analysis of agent software usability is included in Supplementary Note 12.

### Large language model details

ChatDA was evaluated using both GPT-4o, a closed-source model, and Llama 3.3 70B, an open-source model, as its underlying LLM, with model temperatures set to 0.1 to allow for variability in responses. OpenAI Advanced Data Analysis, Open Interpreter, and Data Interpreter were all evaluated with GPT-4o as their underlying LLM. ChatDA, Open Interpreter, and Data Interpreter were assessed systematically using Python scripts, while OpenAI Advanced Data Analysis was evaluated by manually uploading datasets to ChatGPT and obtaining responses through direct queries. To standardize the output for scoring, we used GPT-4o-mini to convert unstructured agent responses into structured formats. Additional implementation details, including system prompts and pre-analysis instructions, are available in Supplementary Notes 6-9.

### Case study: clinical decision support

To assess ChatDA’s practical utility in a real-world setting, we applied it to a proprietary clinical dataset comprising 1,419 knee arthroplasty patients. We investigated whether patient-specific anatomical and functional features could predict the surgeon’s choice between Total Knee Arthroplasty (TKA) and Unicompartmental Knee Arthroplasty (UKA)—a critical decision in knee joint reconstruction. The dataset contained 36 variables, including demographic characteristics (e.g., age, sex, height, and weight), detailed quantitative assessments of knee joint morphology (e.g., bone dimensions, joint angles, and alignment metrics), functional range-of-motion measurements (e.g., flexion and rotation), and structural deformity indicators (e.g., varus/valgus alignment, joint line convergence, and tibial slope irregularities). These features provide a comprehensive characterization of knee joint anatomy, biomechanics, and pathological severity, offering key insights into surgical decision-making. A subset of patients was manually annotated by a fellowship-trained orthopedic surgeon to establish ground truth labels for TKA and UKA. Dataset metadata is provided in Supplementary Note 10.

The primary objective of this analysis was to evaluate whether ChatDA could derive clinically meaningful insights from these features, particularly in predicting surgical decision patterns. Given that many clinicians lack the technical expertise required for advanced statistical modeling, we aimed to assess ChatDA’s ability to bridge this gap by enabling conversational data analysis. Prior to analysis, we ensured compliance with HIPAA Safe Harbor de-identification guidelines (https://www.hhs.gov/hipaa/for-professionals/special-topics/de-identification/index.html)^53^. To further reduce the likelihood of re-identification, we disabled ChatDA’s Python interpreter (tools-only analysis mode), eliminating the LLM’s ability to access individual-level data. This constraint ensured that the LLM processed only structured tool outputs, which were explicitly designed to provide insights at the population level while excluding any patient-specific information (Figure 2A). To simulate real-world clinical use, all queries were phrased in non-technical language, requiring no prior programming or statistical expertise. The full conversational session was completed without manual intervention and is available in Supplementary Note 11.

### Ethics statement

The clinical dataset analyzed in this study was derived from preoperative imaging data originally collected for a previously published study^29^. The corresponding author of that study shared the data with our team. The study from which the data originated had been determined to be exempt from IRB review by Brigham and Women’s Hospital. Our team did not interact with human participants, and upon receipt, we de-identified the dataset in accordance with HIPAA Safe Harbor guidelines.

## Author contributions

A.Y. and Y.M. conceptualized the study. A.Y., J.W., and R.Z. developed the software. A.Y., A.M., and R.Z. curated the datasets for agent benchmarking, performed the benchmarking study, and analyzed the results. J.W. and P.R. contributed to the acquisition and labeling of the knee arthroplasty case study dataset as well as the clinical interpretation of the case study findings. A.Y. and Y.M. were the major contributors in writing the manuscript. All authors read and approved the final manuscript.

## Data availability

All benchmarking datasets used in this study are publicly available, with source information provided in the Supplementary Material. Datasets and scripts for downloading or accessing them are also organized on GitHub: https://github.com/ajy25/TableMage-Analysis. The knee arthroplasty dataset analyzed in the case study contains patient information and cannot be publicly shared. However, the de-identified version of the dataset may be made available by the corresponding author upon reasonable request.

## Code availability

TableMage is released as an open-source Python package on the Python Package Index under the permissive BSD-3-Clause license. Chat Data Analyst can be accessed through a submodule of the TableMage package. All experiments in this study were performed using TableMage version 0.1.0a1. The source code for TableMage and Chat Data Analyst is freely available on GitHub (https://github.com/YMa-lab/TableMage). The AI agent data analysis and machine learning benchmarks complete with datasets and queries are also available on GitHub (https://github.com/ajy25/TableMage-Analysis), along with Python scripts to reproduce similar results.

## Acknowledgements

This work was supported by the Brown University Undergraduate Teaching and Research Award to A.J.Y. and the National Science Foundation (NSF) grant IIS-2500960 to Y.M.

## References

1. Reis, J. Evidence-Based Medicine: Era of Big Data. Biomed. J. Sci. Tech. Res. 41, 32901–32904 (2022).

2. Nascimento, I. J. B. do et al. Impact of Big Data Analytics on People’s Health: Overview of Systematic Reviews and Recommendations for Future Studies. J. Med. Internet Res. 23, e27275 (2021).

3. Radenkovic, D., Keogh, S. B. & Maruthappu, M. Data science in modern evidence-based medicine. J. R. Soc. Med. 112, 493–494 (2019).

4. Gigerenzer, G., Gaissmaier, W., Kurz-Milcke, E., Schwartz, L. M. & Woloshin, S. Helping Doctors and Patients Make Sense of Health Statistics. Psychol. Sci. Public Interest 8, 53–96 (2007).

5. Wang, F., Ma, L., Moulton, G., Wang, M. & Zhang, L. Clinician Data Scientists— Preparing for the Future of Medicine in the Digital World. Health Data Sci. 2022, 9832564.

6. Seth, P. et al. Data Science as a Core Competency in Undergraduate Medical Education in the Age of Artificial Intelligence in Health Care. JMIR Med. Educ. 9, e46344 (2023).

7. Tayebi Arasteh, S., et al. Large language models streamline automated machine learning for clinical studies. Nat. Commun. 15, 1603 (2024).

8. Hou, W. & Ji, Z. Assessing GPT-4 for cell type annotation in single-cell RNA-seq analysis. Nat. Methods 21, 1462–1465 (2024).

9. Meng, X. et al. The application of large language models in medicine: A scoping review. iScience 27, 109713 (2024).

10. Wang, D. & Zhang, S. Large language models in medical and healthcare fields: applications, advances, and challenges. Artif. Intell. Rev. 57, 299 (2024).

11. Singhal, K. et al. Large language models encode clinical knowledge. Nature 620, 172– 180 (2023).

12. Woo, J. J. et al. Custom Large Language Models Improve Accuracy: Comparing Retrieval Augmented Generation and Artificial Intelligence Agents to Noncustom Models for Evidence-Based Medicine. Arthrosc. J. Arthrosc. Relat. Surg. 41, 565–573.e6 (2025).

13. Lewis, P. et al. Retrieval-Augmented Generation for Knowledge-Intensive NLP Tasks. in Advances in Neural Information Processing Systems vol. 33 9459–9474 (Curran Associates, Inc., 2020).

14. Parmar, M. et al. LogicBench: Towards Systematic Evaluation of Logical Reasoning Ability of Large Language Models. in Proceedings of the 62nd Annual Meeting of the Association for Computational Linguistics (Volume 1: Long Papers) (eds Ku, L.-W., Martins, A. & Srikumar, V.) 13679–13707 (Association for Computational Linguistics, Bangkok, Thailand, 2024). doi:10.18653/v1/2024.acl-long.739.

15. Jiang, J., Wang, F., Shen, J., Kim, S. & Kim, S. A Survey on Large Language Models for Code Generation. Preprint at 10.48550/arXiv.2406.00515 (2024).

16. Sun, M., et al. LAMBDA: A Large Model Based Data Agent. Preprint at 10.48550/arXiv.2407.17535 (2024).

17. Hong, S., et al. Data Interpreter: An LLM Agent For Data Science. Preprint at 10.48550/arXiv.2402.18679 (2024).

18. Hu, X., et al. InfiAgent-DABench: Evaluating Agents on Data Analysis Tasks. in Proceedings of the 41st International Conference on Machine Learning 19544–19572 (PMLR, 2024).

19. Aljohani, M., Hou, J., Kommu, S. & Wang, X. A Comprehensive Survey on the Trustworthiness of Large Language Models in Healthcare. Preprint at 10.48550/arXiv.2502.15871 (2025).

20. Atreya, R. V., Smith, J. C., McCoy, A. B., Malin, B. & Miller, R. A. Reducing patient re-identification risk for laboratory results within research datasets. J. Am. Med. Inform. Assoc. JAMIA 20, 95–101 (2013).

21. Sarkar, A. R., Chuang, Y.-S., Mohammed, N. & Jiang, X. De-identification is not enough: a comparison between de-identified and synthetic clinical notes. Sci. Rep. 14, 29669 (2024).

22. Bran, A. M., et al. ChemCrow: Augmenting large-language models with chemistry tools. Preprint at 10.48550/arXiv.2304.05376 (2023).

23. Nakano, R., et al. WebGPT: Browser-assisted question-answering with human feedback. Preprint at 10.48550/arXiv.2112.09332 (2022).

24. Lyu, B., et al. GitAgent: Facilitating Autonomous Agent with GitHub by Tool Extension. Preprint at 10.48550/arXiv.2312.17294 (2023).

25. Parisi, A., Zhao, Y. & Fiedel, N. TALM: Tool Augmented Language Models. Preprint at 10.48550/arXiv.2205.12255 (2022).

26. Campbell, Q., Cox, S., Medina, J., Watterson, B. & White, A. D. MDCrow: Automating Molecular Dynamics Workflows with Large Language Models. Preprint at 10.48550/arXiv.2502.09565 (2025).

27. Lu, P., et al. Chameleon: Plug-and-Play Compositional Reasoning with Large Language Models. Preprint at 10.48550/arXiv.2304.09842 (2023).

28. Sun, M. et al. LAMBDA: A Large Model Based Data Agent. J. Am. Stat. Assoc. 0, 1–13.

29. Woo, J. J. et al. Who Are the Anatomic Outliers Undergoing Total Knee Arthroplasty? A Computed Tomography–Based Analysis of the Hip-Knee-Ankle Axis Across 1,352 Preoperative Computed Tomographies Using a Deep Learning and Computer Vision– Based Pipeline. J. Arthroplasty 39, S188–S199 (2024).

30. Kursa, M. B. & Rudnicki, W. R. Feature Selection with the Boruta Package. J. Stat. Softw. 36, 1–13 (2010).

31. Chen, Y. & Esmaeilzadeh, P. Generative AI in Medical Practice: In-Depth Exploration of Privacy and Security Challenges. J. Med. Internet Res. 26, e53008 (2024).

32. Simon, G. E. et al. Assessing and Minimizing Re-identification Risk in Research Data Derived from Health Care Records. eGEMs 7, 6.

33. Touvron, H., et al. LLaMA: Open and Efficient Foundation Language Models. Preprint at 10.48550/arXiv.2302.13971 (2023).

34. Grattafiori, A., et al. The Llama 3 Herd of Models. Preprint at 10.48550/arXiv.2407.21783 (2024).

35. Care, I. of M. (US) R. on V. & S.-D. H. Healthcare Data as a Public Good: Privacy and Security. in Clinical Data as the Basic Staple of Health Learning: Creating and Protecting a Public Good: Workshop Summary (National Academies Press (US), 2010).

36. Smeltzer, M. P. & Ray, M. A. Statistical considerations for outcomes in clinical research: A review of common data types and methodology. Exp. Biol. Med. 247, 734–742 (2022).

37. Narayan, S. M., Kohli, N. & Martin, M. M. Addressing contemporary threats in anonymised healthcare data using privacy engineering. NPJ Digit. Med. 8, 145 (2025).

38. Rocher, L., Hendrickx, J. M. & de Montjoye, Y.-A. Estimating the success of re-identifications in incomplete datasets using generative models. Nat. Commun. 10, 3069 (2019).

39. Mao, Y., Stevanoski, B. & Montjoye, Y.-A. de. DeSIA: Attribute Inference Attacks Against Limited Fixed Aggregate Statistics. Preprint at 10.48550/arXiv.2504.18497 (2025).

40. Escobar, F. A., Canard, S., Laguillaumie, F. & Phan, D. H. Computational Differential Privacy for Encrypted Databases Supporting Linear Queries. Proc. Priv. Enhancing Technol. (2024).

41. Naveed, M., Kamara, S. & Wright, C. V. Inference Attacks on Property-Preserving Encrypted Databases. in Proceedings of the 22nd ACM SIGSAC Conference on Computer and Communications Security 644–655 (Association for Computing Machinery, New York, NY, USA, 2015). doi:10.1145/2810103.2813651.

42. Harris, C. R. et al. Array programming with NumPy. Nature 585, 357–362 (2020).

43. Mckinney, W. pandas: a Foundational Python Library for Data Analysis and Statistics. Python High Perform. Sci. Comput. (2011).

44. Virtanen, P. et al. SciPy 1.0: fundamental algorithms for scientific computing in Python. Nat. Methods 17, 261–272 (2020).

45. Seabold, S. & Perktold, J. Statsmodels: Econometric and Statistical Modeling with Python. scipy (2010) doi:10.25080/Majora-92bf1922-011.

46. Matplotlib: A 2D Graphics Environment | IEEE Journals & Magazine | IEEE Xplore. https://ieeexplore.ieee.org/document/4160265.

47. Waskom, M. L. seaborn: statistical data visualization. J. Open Source Softw. 6, 3021 (2021).

48. Pedregosa, F. et al. Scikit-learn: Machine Learning in Python. J Mach Learn Res 12, 2825–2830 (2011).

49. Akiba, T., Sano, S., Yanase, T., Ohta, T. & Koyama, M. Optuna: A Next-generation Hyperparameter Optimization Framework. in Proceedings of the 25th ACM SIGKDD International Conference on Knowledge Discovery & Data Mining 2623–2631 (ACM, Anchorage AK USA, 2019). doi:10.1145/3292500.3330701.

50. Liu, J. LlamaIndex. (2022).

51. Grinsztajn, L., Oyallon, E. & Varoquaux, G. Why do tree-based models still outperform deep learning on typical tabular data? in *Proceedings of the 36th International Conference on Neural Information Processing Systems* 507–520 (Curran Associates Inc., Red Hook, NY, USA, 2022).

52. Vanschoren, J., van Rijn, J. N., Bischl, B. & Torgo, L. OpenML: networked science in machine learning. SIGKDD Explor Newsl 15, 49–60 (2014).

53. Rights (OCR), O. for C. Guidance Regarding Methods for De-identification of Protected Health Information in Accordance with the Health Insurance Portability and Accountability Act (HIPAA) Privacy Rule. https://www.hhs.gov/hipaa/for- professionals/special-topics/de-identification/index.html (2012).

