## Supplementary Material for "Tool-wielding language-model-based agent offers conversational exploration of clinical tabular data"

#### Table of Contents


**SUPPLEMENTARY FIGURES** ... **3**

Supplementary Figure 1. Data analysis benchmark (DataAnalysisQA) results. ... 3
Supplementary Figure 2. Machine learning regression benchmark results. ... 4
Supplementary Figure 3. Machine learning classification benchmark results. ... 5
Supplementary Figure 4. DataAnalysisQA failure analysis. ... 6
Supplementary Figure 5. DataAnalysisQA ChatDA tool-level error rate. ... 7

**SUPPLEMENTARY TABLES** ... **9**

Supplementary Table 1. Table of ChatDA tools. ... 9
Supplementary Table 2. DataAnalysisQA dataset details. ... 10
Supplementary Table 3. Machine learning benchmark task details. ... 11
Supplementary Table 4. Summary of proprietary dataset used in case study. ... 12
Supplementary Table 5. Comparison between clinical trial management systems platforms and ChatDA. ... 14

**SUPPLEMENTARY NOTES** ... **15**

Supplementary Note 1. Additional discussion: ChatDA data privacy considerations and limitations. ... 15
*Reducing re-identification risk for de-identified data* ... *15*
*Limitations and residual risks* ... *15*
*Practical guidance* ... *16*
Supplementary Note 2. Additional discussion: Limitations of ChatDA. ... 17
Supplementary Note 3. Additional discussion: Limitations of the agent benchmarking methodology. ... 18
Supplementary Note 4. Brief investigation of prevalence of tabular data in clinical research. ... 19
Supplementary Note 5. InfiAgent-DABench results. ... 20
Supplementary Note 6. Large language model details. ... 21
Supplementary Note 7. Competing agent details ... 22
Supplementary Note 8. DataAnalysisQA benchmark prompts and templates. ... 23
Supplementary Note 9. Machine learning benchmark prompts and templates ... 25
Supplementary Note 10. Clinical case study knee arthroplasty dataset details. ... 26
Supplementary Note 11. Clinical case study conversation transcript. ... 27
Supplementary Note 12. Agent software usability. ... 37

**REFERENCES** ... **38**

#### Supplementary Figures

**Supplementary Figure 1. Data analysis benchmark (DataAnalysisQA) results.**

**(A)** Bar plot depicting the accuracy of Chat Data Analyst (ChatDA) powered by GPT-4o (dark purple), ChatDA powered by GPT-4o with the tools-only restriction (purple), ChatDA powered by Llama 3.3 70B (light purple), OpenAI Advanced Data Analysis (orange), Data Interpreter (green), Open Interpreter (blue), and LAMBDA (turquoise) on the 100-question DataAnalysisQA benchmark. Accuracies stratified by question category are also reported. **(B)** Same as panel (A), but with indexing-related questions excluded. Non-coding agents, such as ChatDA operating in tools-only mode, exhibit inherent limitations in handling data analysis tasks that involve indexing operations. Excluding indexing questions allows for a more accurate assessment of non-coding agent performance.

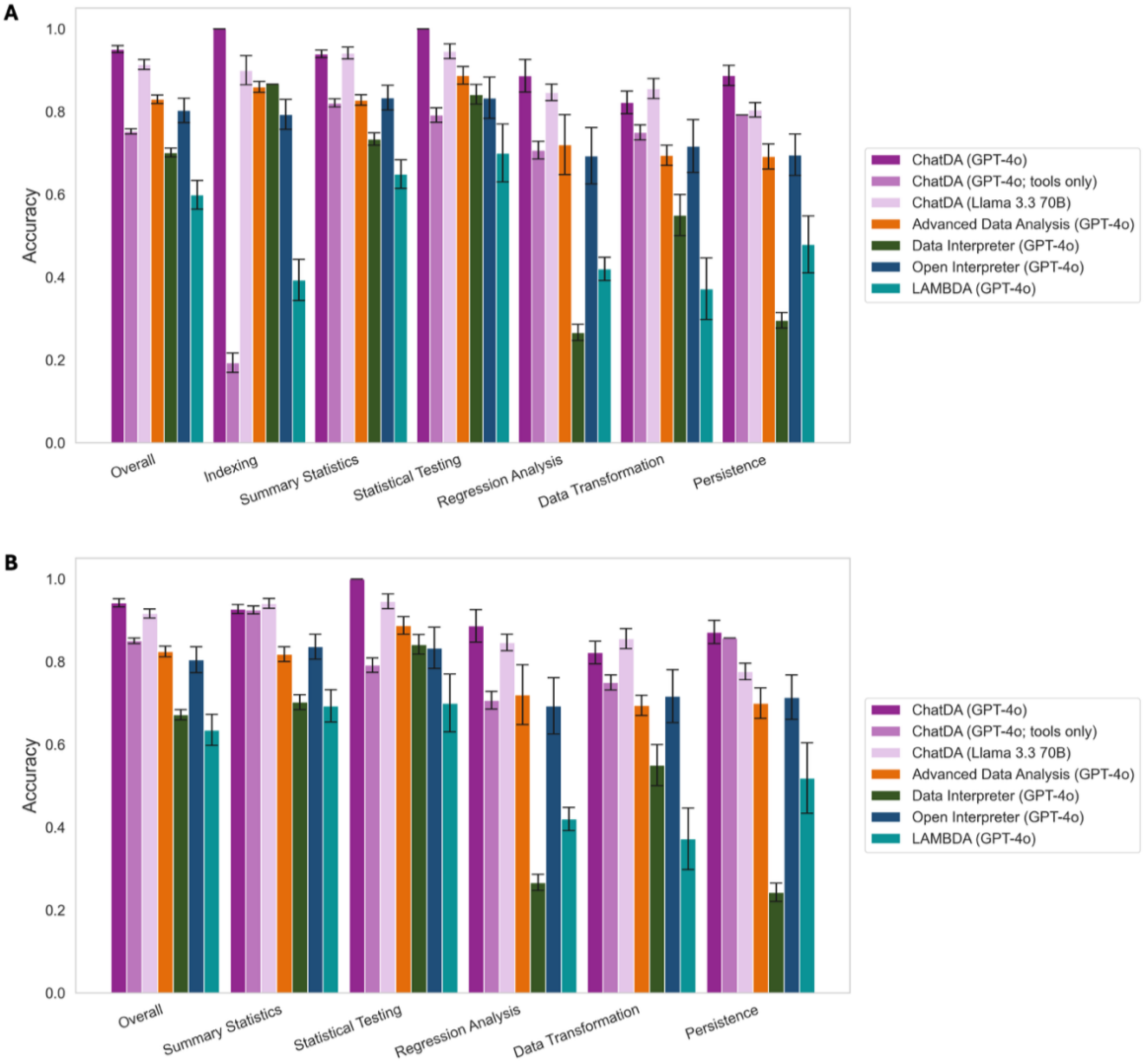

**Supplementary Figure 2. Machine learning regression benchmark results.**

Machine learning regression benchmark results for Chat Data Analyst (ChatDA) powered by GPT-4o (dark purple), ChatDA powered by Llama 3.3 70B (light purple), OpenAI Advanced Data Analysis (orange), Data Interpreter (green), Open Interpreter (blue), and LAMBDA (turquoise). 95% confidence intervals of the root mean squared errors (RMSE; lower is better) generated by the agents across 10 replicates are reported in the legend. Each task is identified by its unique OpenML Task ID.

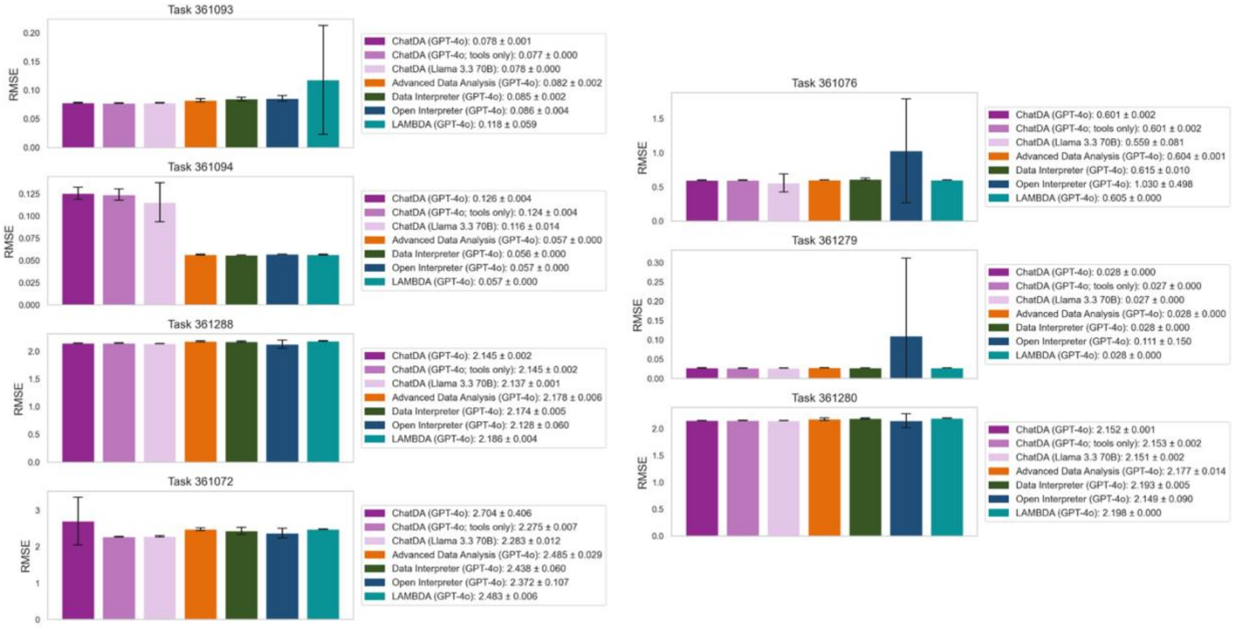

**Supplementary Figure 3. Machine learning classification benchmark results.**

Machine learning classification benchmark results for Chat Data Analyst (ChatDA) powered by GPT-4o (dark purple), ChatDA powered by Llama 3.3 70B (light purple), OpenAI Advanced Data Analysis (orange), Data Interpreter (green), Open Interpreter (blue), and LAMBDA (turquoise). 95% confidence intervals of the area under the receiver operating characteristic curve (AUC; higher is better) generated by the agents across 10 replicates are reported in the legend. Each task is identified by its unique OpenML Task ID.

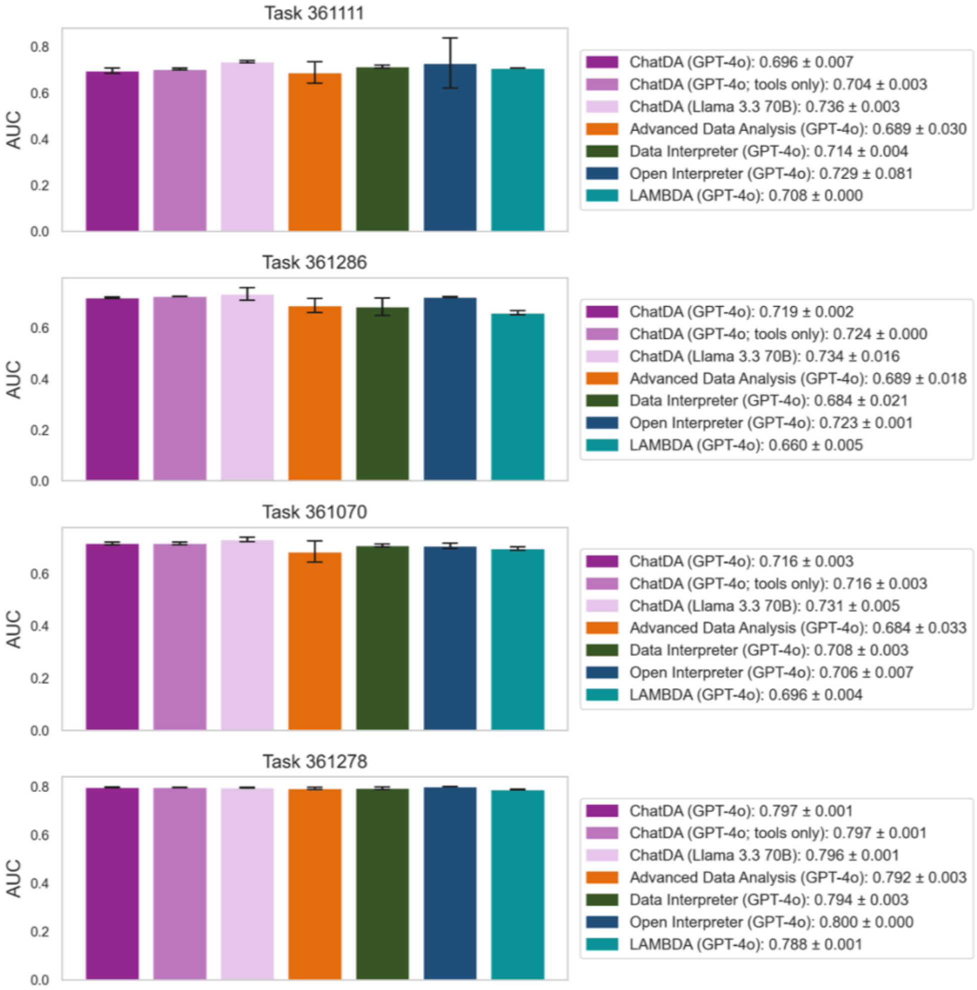

**Supplementary Figure 4. DataAnalysisQA failure analysis.**

**(A)** Bar plot depicting the failure rate (Methods) of Chat Data Analyst (ChatDA) powered by GPT-4o (dark purple), ChatDA powered by GPT-4o with the tools-only restriction (purple), ChatDA powered by Llama 3.3 70B (light purple), OpenAI Advanced Data Analysis (orange), Data Interpreter (green), Open Interpreter (blue), and LAMBDA (turquoise) on the 100-question DataAnalysisQA benchmark. Failure rates stratified by question category are also reported. **(B)** Same as panel (A), but with indexing-related questions excluded.

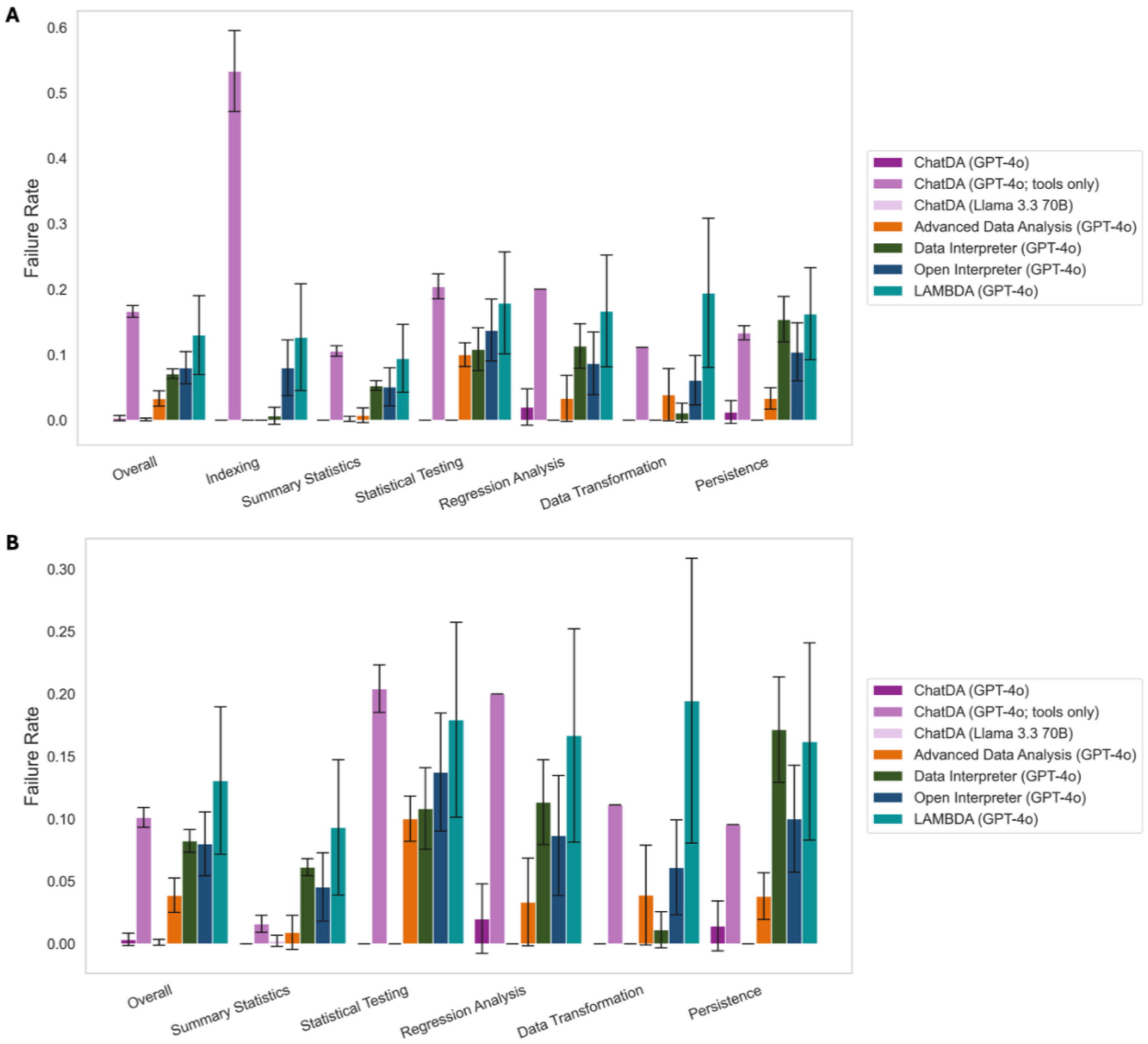

**Supplementary Figure 5. DataAnalysisQA ChatDA tool-level error rate.**

ChatDA accuracy on the DataAnalysisQA benchmark stratified by expected tool use (Methods) for **(A)** ChatDA (GPT-4o), **(B)** ChatDA (GPT-4o; tools only), and **(C)** ChatDA (Llama 3.3 70B).

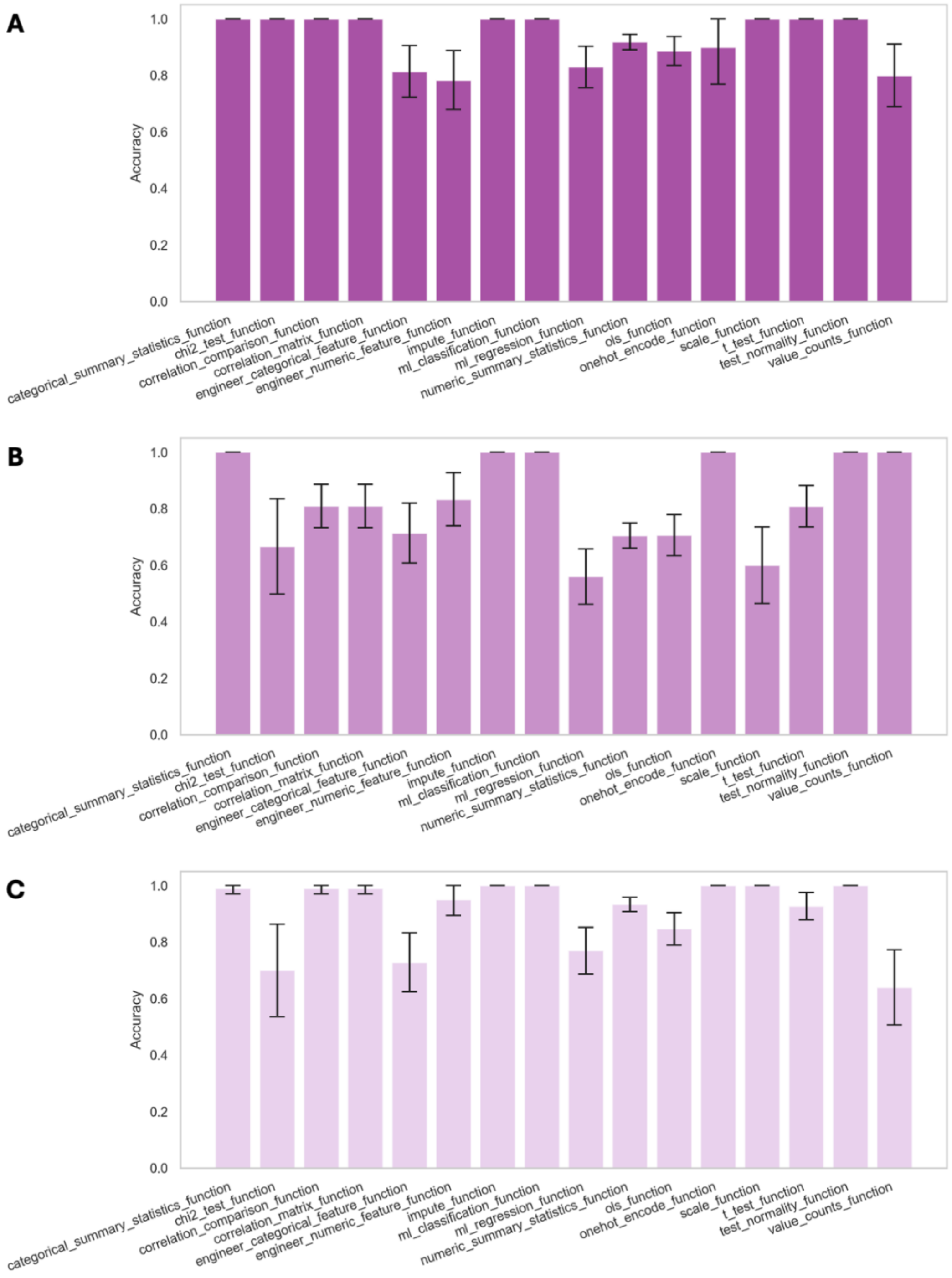

**Supplementary Figure 6. Prevalence of tabular data in clinical research.**

Prevalence of tabular versus non-tabular data in NEJM research articles, 2005-2025. **(A)** MeSH-based filtering of all NEJM clinical research articles indexed in PubMed (n = 30,416) yielded 3,425 full-length studies with clinical data. Of these, 3,181 (92.9%) were non-imaging and therefore primarily tabular, while 244 (7.1%) involved imaging modalities. **(B)** Independent random sample of 2,000 NEJM articles between 2005-2025 identified 334 full-length clinical studies. Of these, 253 (75.7%) contained exclusively tabular data, while 81 (24.3%) incorporated non-tabular modalities.

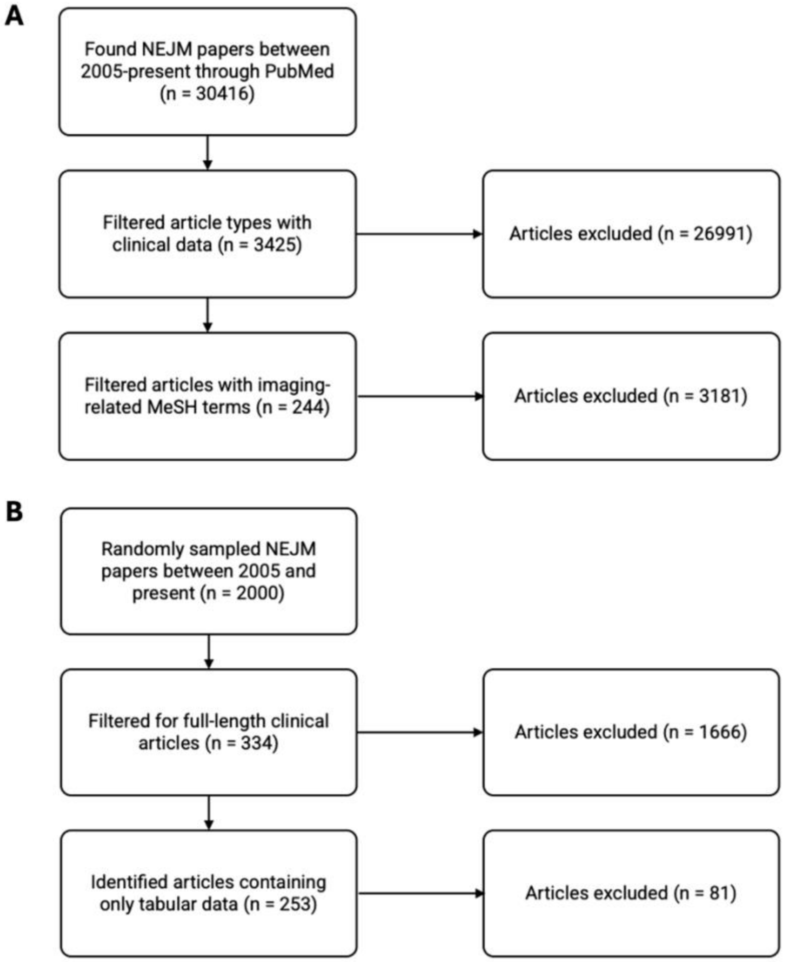

### Supplementary Tables

#### Supplementary Table 1. Table of ChatDA tools.

Table listing all custom tools defined for ChatDA's use. For each query, only the top *k* number of tools most relevant to the query, as determined by retrieval-augmented generation on the tools' metadata, are made available to ChatDA's underlying LLM.

| *Category* | *Tool/Method Name* | *Description* |
|---|---|---|
| ***Exploratory Data Analysis*** | numeric_summary_statistics_function | Computes summary statistics for the numeric variables. |
| | categorical_summary_statistics_function | Computes summary statistics for the categorical variables. |
| | correlation_comparison_function | Calculates the correlation between a single numeric variable and each variable in each list of numeric variables. |
| | correlation_matrix_function | Calculates the correlation matrix for a list of numeric variables. |
| | chi2_test_function | Conducts a Chi-square test to assess the independence between two categorical variables. |
| | t_test_function | Conducts a t-test. Supports Student's, Welch's, Yuen's, and Mann-Whitney variations. |
| | anova_test_function | Conducts a one-way ANOVA. Supports the standard one-way ANOVA and the Kruskal-Wallis variation. |
| | test_normality_function | Tests for the normality of a numeric variable. Supports Shapiro-Wilk, Kolmogorov-Smirnov, or Anderson-Darling tests. |
| | plot_function | Visualizes the distribution of a single variable using a histogram or bar plot or explores the relationship between two variables with options for a scatterplot, box plot, or crosstab heatmap. |
| | plot_pairs_function | Creates a grid of visualizations to explore pairwise relationships among a list of variables, |
| | value_counts_function | Counts the number of occurrences for each unique value of a variable. |
| ***Data Transformation*** | engineer_numeric_feature_function | Engineers a new numeric variable based on a formula of numeric variables. |
| | engineer_categorical_feature_function | Engineers a new categorical variable from a numeric variable based on a list of thresholds. |
| | scale_function | Scales numeric variables. Supports standard, min-max, log, and log1p transformations. |
| | impute_function | Imputes missing numeric and categorical variable values. Supports mean, median, and k-nearest neighbors for numeric variables. Supports most frequent or new level for categorical variables. |
| | drop_na_function | Drops all rows with missing values. |
| | drop_highly_missing_vars_function | Drops columns that are highly missing, as defined by a threshold. |
| | onehot_encode_function | One-hot encodes specified categorical variables. |
| ***Causal Inference*** | estimate_causal_effect_function | Estimates the ATE or ATT. Supports outcome regression, IPW-weighted regression, and IPW estimator. |
| ***Regression Analysis*** | ols_function | Ordinary least squares regression. Supports stepwise selection and nested model testing. |
| | logit_function | Logistic regression. Supports stepwise selection. |
| ***Machine Learning*** | ml_regression_function | Machine learning regressor benchmarking. |
| | ml_classification_function | Machine learning classifier benchmarking. |
| | clustering_function | Clustering visualization. |
| | feature_selection_function | Feature selection. Supports voting selection for more than one feature selection method. |

**Supplementary Table 2. DataAnalysisQA dataset details.**

Table listing all datasets used in the DataAnalysisQA benchmark along with their sources, their shapes, and the number of benchmark questions for each dataset. The CSV files for each dataset are organized along with the DataAnalysisQA questions, available at https://github.com/ajy25/TableMage-Analysis.

| ***Dataset Name*** | ***Dataset Source*** | ***Example Count*** | ***Feature Count*** | ***Question Count*** |
|---|---|---|---|---|
| ***Breast Cancer*** | scikit-learn (https://scikit-learn.org/stable/modules/generated/sklearn.datasets.load_breast_cancer.html) | 569 | 31 | 9 |
| ***Credit*** | InfiAgent-DABench (https://github.com/InfiAgent/InfiAgent) | 400 | 11 | 10 |
| ***House Prices*** | Kaggle (https://www.kaggle.com/competitions/house-prices-advanced-regression-techniques/data) | 1460 | 80 | 12 |
| ***Titanic*** | Kaggle (https://www.kaggle.com/competitions/titanic/data) | 891 | 12 | 10 |
| ***Wine*** | scikit-learn (https://scikit-learn.org/stable/modules/generated/sklearn.datasets.load_wine.html) | 178 | 14 | 9 |
| ***Abalone*** | InfiAgent-DABench (https://github.com/InfiAgent/InfiAgent) | 4177 | 9 | 11 |
| ***Baseball*** | InfiAgent-DABench (https://github.com/InfiAgent/InfiAgent) | 337 | 17 | 10 |
| ***Auto MPG*** | InfiAgent-DABench (https://github.com/InfiAgent/InfiAgent) | 392 | 8 | 10 |
| ***Healthcare*** | Kaggle (https://www.kaggle.com/datasets/prasad22/healthcare-dataset) | 55500 | 15 | 10 |
| ***Iris*** | Kaggle (https://www.kaggle.com/datasets/uciml/iris) | 150 | 6 | 9 |

**Supplementary Table 3. Machine learning benchmark task details.**

Table listing OpenML task IDs used in the machine learning benchmark along with relevant metadata. The tasks comprise the subset of the tabular dataset machine learning benchmark curated by Grinsztajn et al.[1] with 10,000 or fewer rows and 100 or fewer columns. The CSV files for the datasets, along with a Python script to obtain the datasets from OpenML, are available at https://github.com/ajy25/TableMage-Analysis.

| *OpenML Task ID* | *Dataset Name* | *Task Type* | *Feature Datatypes* | *Target Variable* | *Example Count* | *Feature Count* |
|---|---|---|---|---|---|---|
| ***361111*** | eye_movements | classification | numeric and categorical | label | 7608 | 23 |
| ***361286*** | compas-two-years | classification | numeric and categorical | twoyearrecid | 4966 | 11 |
| ***361070*** | eye_movements | classification | numeric only | label | 7608 | 20 |
| ***361278*** | heloc | classification | numeric only | RiskPerformance | 10000 | 22 |
| ***361093*** | analcatdata_supreme | regression | numeric and categorical | Log_exposure | 4052 | 7 |
| ***361094*** | visualizing_soil | regression | numeric and categorical | track | 8641 | 4 |
| ***361288*** | abalone | regression | numeric and categorical | Classnumberofrings | 4177 | 8 |
| ***361072*** | cpu_act | regression | numeric only | usr | 8192 | 21 |
| ***361076*** | wine_quality | regression | numeric only | quality | 6497 | 11 |
| ***361279*** | yprop_4_1 | regression | numeric only | oz252 | 8885 | 42 |
| ***361280*** | abalone | regression | numeric only | Classnumberofrings | 4177 | 7 |

**Supplementary Table 4. Summary of proprietary dataset used in case study.**

Table containing summary statistics for variables included in the knee arthroplasty case study dataset.

| | | Grouped by Arthroplasty Classification | | | |
|---|---|---|---|---|---|
| | | **Missing** | **Overall** | **TKA** | **UKA** |
| **n** | | | 1419 | 471 | 324 |
| **Age, mean (SD)** | | 7 | 67.3 (8.6) | 67.5 (8.8) | 66.8 (8.3) |
| **Laterality, n (%)** | **left** | | 709 (50.0) | 239 (50.7) | 149 (46.0) |
| | **right** | | 706 (49.8) | 232 (49.3) | 175 (54.0) |
| **Height, mean (SD)** | | 1163 | 1.7 (0.1) | 1.7 (0.1) | 1.7 (0.1) |
| **Weight, mean (SD)** | | 107 | 85.7 (17.3) | 84.9 (17.9) | 86.7 (17.1) |
| **Sulcus Angle, mean (SD)** | | 84 | 135.6 (8.7) | 135.7 (8.5) | 135.9 (8.3) |
| **Trochlear Groove Width, mean (SD)** | | 84 | 36.7 (4.5) | 36.3 (4.5) | 36.6 (4.2) |
| **Posterior Condylar Angle, mean (SD)** | | 84 | 7.4 (5.7) | 7.5 (6.0) | 7.2 (5.7) |
| **TEA Horizontal Angle, mean (SD)** | | 84 | 7.4 (5.8) | 7.7 (6.0) | 7.7 (5.9) |
| **TEA-PCA Angle, mean (SD)** | | 84 | 5.0 (2.3) | 5.1 (2.3) | 5.0 (2.3) |
| **TT-TG Distance, mean (SD)** | | 87 | 12.0 (4.9) | 12.6 (5.2) | 11.6 (4.6) |
| **Medial Posterior Tibial Slope, mean (SD)** | | 84 | 4.9 (4.3) | 5.2 (4.6) | 4.8 (4.0) |
| **Lateral Posterior Tibial Slope, mean (SD)** | | 84 | 4.4 (4.3) | 4.0 (3.9) | 3.6 (3.7) |
| **AP Femur Distance, mean (SD)** | | 87 | 60.1 (5.7) | 60.8 (5.6) | 60.5 (4.8) |
| **AP Tibial Distance, mean (SD)** | | 85 | 50.0 (5.0) | 50.5 (5.0) | 50.0 (4.8) |
| **External Rotation, n (%)** | **False** | | 798 (56.2) | 274 (58.2) | 182 (56.2) |
| | **None** | | 109 (7.7) | 7 (1.5) | 9 (2.8) |
| | **True** | | 512 (36.1) | 190 (40.3) | 133 (41.0) |
| **Knee Flexion Angle, mean (SD)** | | 107 | 7.8 (7.5) | 7.8 (6.3) | 7.4 (5.6) |
| **Femoral Neck Angle, mean (SD)** | | 99 | 9.5 (9.6) | 9.6 (9.2) | 10.8 (9.1) |
| **Neck-Shaft Angle, mean (SD)** | | 80 | 128.3 (4.5) | 128.5 (4.3) | 128.7 (4.7) |
| **Femoral Anteversion, mean (SD)** | | 106 | 11.5 (9.4) | 11.4 (9.4) | 11.8 (9.7) |
| **Femoral Torsion, mean (SD)** | | 112 | 12.5 (7.6) | 12.6 (7.5) | 12.8 (7.8) |
| **Acetabular Angle, mean (SD)** | | 92 | 8.4 (7.5) | 7.0 (7.4) | 7.7 (7.1) |
| **Femoral Head Width, mean (SD)** | | 133 | 44.2 (4.2) | 44.1 (4.1) | 44.5 (4.0) |
| **Joint Line Convergence Angle, mean (SD)** | | 104 | 2.7 (1.8) | 2.5 (1.8) | 3.0 (1.7) |
| **Hip-Knee-Ankle Angle, mean (SD)** | | 85 | 173.5 (4.2) | 173.7 (4.2) | 173.0 (4.3) |
| **Lateral Distal Femoral Angle, mean (SD)** | | 81 | 87.9 (3.5) | 87.6 (3.7) | 88.4 (3.3) |

| | | | | | |
|---|---|---|---|---|---|
| **Medial Proximal Tibial Angle, mean (SD)** | | 84 | 85.9 (3.8) | 85.9 (4.0) | 85.3 (3.5) |
| **Varus Alignment, n (%)** | **False** | | 359 (25.3) | 157 (33.3) | 56 (17.3) |
| | **None** | | 78 (5.5) | 5 (1.1) | 3 (0.9) |
| | **True** | | 982 (69.2) | 309 (65.6) | 265 (81.8) |
| **Tibiofemoral Angle, mean (SD)** | | 79 | 4.5 (4.0) | 5.3 (4.5) | 3.8 (3.6) |
| **Patella Width, mean (SD)** | | 89 | 44.0 (4.2) | 44.0 (4.1) | 43.9 (4.1) |
| **Medial Femoral Condyle Width, mean (SD)** | | 88 | 28.1 (3.0) | 28.2 (3.1) | 28.3 (2.9) |
| **Lateral Femoral Condyle Width, mean (SD)** | | 91 | 29.0 (3.1) | 28.9 (2.8) | 28.8 (3.0) |
| **Tibial Width, mean (SD)** | | 93 | 76.6 (6.5) | 76.9 (6.6) | 76.6 (6.1) |
| **Medial Tibial Width, mean (SD)** | | 92 | 29.2 (3.4) | 29.0 (3.3) | 29.1 (3.2) |
| **Lateral Tibial Width, mean (SD)** | | 92 | 29.9 (3.5) | 30.0 (3.6) | 29.5 (3.1) |
| **Sex, n (%)** | **F** | | 805 (56.7) | 282 (59.9) | 174 (53.7) |
| | **M** | | 614 (43.3) | 189 (40.1) | 150 (46.3) |

**Supplementary Table 5. Comparison between clinical trial management systems platforms and ChatDA.**

Table summarizing key differences in purpose, functionality, and user interaction between traditional clinical data platforms (here, we focus on REDCap[2], a clinical trial management system (CTMS) and electronic data capture platform) and ChatDA. While REDCap focus on secure data capture, management, and compliance, ChatDA provides natural language-driven analytic capabilities for de-identified tabular datasets. The systems are therefore complementary: traditional platforms ensure clinical data integrity, while ChatDA enables downstream statistical analysis, machine learning, and exploratory interpretation of de-identified tables.

| Feature | CTMS (REDCap) | ChatDA |
|---|---|---|
| Primary purpose | Secure data capture, project management, and regulatory compliance | Analytic workspace for de-identified datasets, supporting exploratory and advanced analysis |
| Core functions | Data entry, storage, workflow management, participant de-identification | Natural language-driven statistical analysis, machine learning, visualization |
| User interaction | Data upload, form-based collection, structured project setup | Conversational querying and analysis of user-provided data |
| Positioning | Foundational infrastructure ensuring data integrity and governance | AI system focused on data analysis rather than data management |

### Supplementary Notes

**Supplementary Note 1. Additional discussion: ChatDA data privacy considerations and limitations.**

*Reducing re-identification risk for de-identified data*

A core concern with AI-assisted data analysis is inadvertent exposure of sensitive information to external cloud providers. Even when datasets are de-identified under standards such as HIPAA Safe Harbor, disclosure of detailed, record-level information to a third-party service can still increase the risk of re-identification through linkage or inference.

Conventional cloud-based code-writing agents amplify this risk. Because they operate by generating and executing arbitrary code, they can access individual records, extract detailed subsets, or produce intermediate outputs (e.g., row extracts, debugging logs, or patient-level predictions) that exceed the protections afforded by de-identification. In practice, this means that even legally de-identified datasets may become vulnerable if analyzed through unrestricted cloud LLM agents.

ChatDA's "tools-only" mode addresses this gap by structurally limiting the LLM's role. In this mode:

- All computations are executed locally. Analytic operations are performed within the user's environment, not in the cloud.
- The LLM cannot index or inspect records. The underlying LLM is never provided with individual-level observations.
- Only aggregated outputs are shared. The LLM receives population-level summaries, such as descriptive statistics, regression coefficients, test results, and model performance metrics.

This architecture prevents direct exposure of records while retaining the usability of conversational AI. By design, the outputs align with standard practices in clinical research, where individual-level disclosures are avoided. In this way, ChatDA enables practical analysis of de-identified data with a lower residual risk profile than conventional code-writing agents, while still providing clinicians with accessible analytical support.

*Limitations and residual risks*

The "tools-only" mode should be understood as a risk-mitigation mechanism, not as a complete privacy solution. Several limitations remain:

1. Re-identification from aggregates. Modern privacy research shows that releasing multiple summary statistics over time can enable inference attacks. Even without

direct identifiers, small subgroup counts, extreme values, or repeated queries can make individuals more identifiable[3].

2. Cumulative disclosure risk. Outputs from separate queries may be combined to reconstruct sensitive attributes. Over many interactions, the accumulation of population-level statistics can erode the protection of de-identification[4].
3. Small or rare subgroups. When the dataset includes small cohorts (e.g., rare diseases, specific demographic intersections), summary statistics may still reveal membership in those groups. This remains a significant concern for clinical datasets with imbalanced representation[5].
4. Statistical disclosure through modeling. Advanced inference techniques can exploit regression coefficients, p-values, or feature importances to infer sensitive properties of individuals, especially when the adversary has auxiliary datasets[6].
5. No formal privacy guarantees. Unlike differential privacy or cryptographic approaches[7], ChatDA does not provide mathematical guarantees against disclosure. It reduces—but does not eliminate—re-identification risk.

*Practical guidance*

ChatDA is intended for use with datasets that have been properly de-identified in accordance with established regional standards such as the HIPAA Safe Harbor method (https://www.hhs.gov/hipaa/for-professionals/special-topics/de-identification/index.html). In practice, this means ChatDA should only be applied to datasets that can legally be analyzed and stored using cloud resources. In the United States, under HIPAA, properly de-identified data is not considered protected health information (PHI). Other regions impose additional standards, such as the California Consumer Privacy Act (https://oag.ca.gov/privacy/ccpa) and the European Union General Data Protection Regulation (https://gdpr-info.eu/).

When operating on data de-identified according to these legal frameworks, ChatDA in “tools-only” mode reduces the risk of exposing individual-level information to external providers. We present ChatDA as a lower-risk alternative for analyzing de-identified clinical data compared to existing code-generating agents.

However, de-identification is not equivalent to anonymity. Residual risks of re-identification through statistical inference remain, particularly for small or unique subgroups. For high-sensitivity applications, organizations should consider complementary safeguards such as suppressing or perturbing outputs tied to small cell counts, query logging, or formal privacy-preserving methods. In summary, ChatDA can be used responsibly with properly de-identified datasets, but its deployment should be framed as part of a broader privacy risk management strategy rather than as a standalone guarantee of confidentiality.

**Supplementary Note 2. Additional discussion: Limitations of ChatDA.**

A key limitation of this work is that the evaluation of ChatDA was performed exclusively on tabular datasets. While structured, table-formatted data are foundational to many areas of clinical research (Supplementary Note 4), they represent only one modality of clinical data. Modern biomedical research increasingly incorporates imaging, free-text clinical notes, genomics, and multimodal datasets, each of which presents distinct analytic challenges. By restricting scope to tabular data, ChatDA cannot yet address analytic needs for unstructured text (such as clinical narratives), signal data (such as ECG or waveforms), or radiology and pathology imaging. This limitation reduces the system's applicability compared with general-purpose agents that permit free-form code execution across diverse data types[8]. The functionality of ChatDA is further bounded by the predefined tools in its environment, meaning that analyses outside the scope of these tools cannot be performed without extending the toolkit. This limitation in flexibility is evidenced in ChatDA's underperformance on the coding-specific InfiAgent-DABench benchmark (Supplementary Note 5).

We also note that ChatDA does not achieve perfect accuracy on the DataAnalysisQA benchmark (Supplementary Figures 1, 4, and 5). In tools-only mode, accuracy declined relative to full mode, with notable difficulties in applying the regression tool and several statistical testing and transformation tools (Supplementary Figure 5). Tools-only ChatDA also failed more often, being unable to return an answer in a larger fraction of cases (Supplementary Figure 4). Although the interface allows users to monitor and correct outputs, these limitations highlight the fragility of tool-restricted agents. Future work should investigate the causes of such errors and explore methods to improve tool execution. Continued advances in LLMs may reduce these failure rates over time.

#### Supplementary Note 3. Additional discussion: Limitations of the agent benchmarking methodology.

This work has several limitations that should be acknowledged regarding the benchmarking strategy. First, the development of DataAnalysisQA as a new evaluation benchmark inherently introduces bias. Although it was designed to improve upon limitations of a prior framework, InfiAgent-DABench[9] (see Methods), the process of question curation and answer validation inevitably reflects the design choices and assumptions of its creators. This can shape the types of reasoning and operations emphasized and may not generalize fully to other evaluation settings.

Second, while DataAnalysisQA was inspired by InfiAgent-DABench, the majority of questions were intentionally constructed to be solvable with the tools provided by ChatDA. This introduces a structural bias that favors ChatDA's tool-based design. As a result, ChatDA's strong performance on DataAnalysisQA should be interpreted with caution. On alternative benchmarks emphasizing tasks that require unconstrained code generation or tasks for which ChatDA does not have a predefined tool, its performance would likely be lower, since its capabilities depend on function calling rather than free-form program synthesis. Indeed, we verify this hypothesis in Supplementary Note 5 on the Python-specific InfiAgent-DABench data analysis benchmark. More broadly, the functionality of ChatDA is bounded by the breadth of tools available to it, which means that missing tools directly constrain its performance. ChatDA's performance on DataAnalysisQA should therefore be interpreted with caution, as missing tools directly constrain its capabilities and may not generalize to tasks outside its current toolkit.

Third, the machine learning benchmark used here was limited in scope to small- and medium-scale tabular datasets. While sufficient for demonstrating feasibility for tabular data, the benchmark does not encompass more complex ML tasks such as large-scale datasets, multimodal integration, survival analysis, or deep learning architectures. We note that ChatDA does not currently have tooling to perform custom deep learning modeling, multimodal data analysis, or survival analysis (see Supplementary Note 2).

Benchmarking LLM agents remains an unresolved challenge. Current benchmarks risk embedding the biases of their designers and may inadvertently privilege certain model architectures or tool-use paradigms. To advance the field, it will be necessary to develop community-driven efforts that promote standardized, transparent, and paradigm-agnostic benchmarks.

**Supplementary Note 4. Brief investigation of prevalence of tabular data in clinical research.**

Tabular datasets—structured as rows representing patients and columns representing variables—form the backbone of most clinical research. To empirically assess their prevalence, we conducted a structured review of articles published in the New England Journal of Medicine (NEJM) between 2005 and 2025. Using PubMed abstracts and Medical Subject Headings (MeSH), we classified studies according to whether their primary data were tabular (e.g., demographics, laboratory values, outcomes in spreadsheet-like form) or non-tabular (e.g., imaging, waveforms, free text, or genomics). Two complementary approaches were implemented.

First, a random sample of 2000 NEJM articles between 2005-2025 was screened. 334 full-length, original research articles were extracted from this set of articles. Of these, 253 (75.7%) relied exclusively on tabular data, while 81 (24.3%) incorporated non-tabular modalities. Second, we performed MeSH-based classification of all NEJM full-length, original research articles on PubMed between 2005 and 2025, yielding 3425 full-length articles suitable for analysis. Within this larger sample, 3181 articles (92.9%) were non-imaging and therefore primarily tabular, while 244 (7.1%) involved imaging modalities. These results are visualized in Supplementary Figure 6.

Together, these analyses confirm that the majority of clinical research in a leading general medical journal is grounded in tabular data. This prevalence reflects both practicality and methodological tradition: structured patient-level variables are amenable to standard statistical methods, align with reporting requirements for clinical trials and observational studies, and can be readily aggregated for population-level inference. The dominance of tabular data underscores its centrality to evidence-based medicine and justifies the design of specialized tools such as ChatDA and TableMage to support their analysis.

#### Supplementary Note 5. InfiAgent-DABench results.

We compared ChatDA against Data Interpreter[10], Open Interpreter, and LAMBDA[8], on the InfiAgent-DABench data analysis agent benchmark[9]. Although we emphasize the limitations of this benchmark and perform an independent evaluation using DataAnalysisQA (Methods), we report InfiAgent-DABench results here to illustrate ChatDA's relative shortcomings in flexible data analysis. InfiAgent-DABench is, to our knowledge, the only peer-reviewed systematic benchmark of data analysis agents at the time of writing. However, it suffers from several flaws, including reliance on code-specific technical prompts, AI-generated ground truth, and the absence of multi-question continuity for testing persistent transformations (Methods).

On this benchmark, Data Interpreter (0.833), Open Interpreter (0.773), and LAMBDA (0.778) outperformed ChatDA (0.638) in accuracy by question. This result is expected, as ChatDA prioritizes tool-based operations whereas InfiAgent-DABench prioritizes Python code execution. For example, ChatDA failed on question 70, which explicitly required the use of sklearn.linear_model.LinearRegression() with a specified train-test split and RMSE calculation. In contrast, DataAnalysisQA includes questions with more flexible instructions, such as: "Regress 'SalePrice' on 'TotalSF,' 'LotShape,' and 'GarageArea' with linear regression. Report the train R-squared and the test RMSE." Such phrasing aligns better with ChatDA's design, which presupposes a pre-initialized train/test split and does not depend on explicit Python class invocation.

We attribute ChatDA's underperformance on InfiAgent-DABench to two design factors. First, ChatDA is optimized to execute high-level tool calls, not to replicate Python syntax. Benchmark questions referencing specific classes and functions reduce its ability to select the correct operation. Second, ChatDA enforces a pre-initialized train-test split to minimize pipelining errors (Methods). This prevents dynamic re-splitting in response to question-specific instructions, which explains its very low score on machine learning tasks (ChatDA: 0.158; Data Interpreter: 0.684; Open Interpreter: 0.632; LAMBDA: 0.579).

This benchmark also highlights a broader limitation of ChatDA: inflexibility. Unlike code-generating agents that can synthesize arbitrary scripts, ChatDA is constrained by the functions available in its predefined toolkit and by its rigid handling of workflows such as dataset splitting. These constraints make ChatDA more robust in terms of consistency and data privacy but limit its adaptability to benchmarks or tasks framed with code-specific or highly variable requirements. While this trade-off is acceptable in clinical data analysis—where reproducibility, safety, and interpretability are paramount—it reduces ChatDA's competitiveness on benchmarks optimized for unconstrained coding agents.

**Supplementary Note 6. Large language model details.**

GPT-4o (gpt-4o-2024-08-06) inference was provided by OpenAI through the OpenAI Platform (https://platform.openai.com/docs/models#gpt-4o). GPT-4o was used to power ChatDA, Data Interpreter, and Open Interpreter. The February 2025 version of the ChatGPT Plus (https://chatgpt.com) GPT-4o model was used for OpenAI Advanced Data Analysis benchmarking.

ChatDA was also evaluated with Llama 3.3 70B, an open-source Llama model with 70 billion parameters, as its underlying LLM. Llama 3.3 70B can be hosted locally on powerful hardware (https://www.llama.com/docs/llama-everywhere). Due to limited computational resources, Llama 3.3 70B (llama-3.3-70b-versatile) inference was provided by Groq Cloud (https://console.groq.com) for this study.

#### Supplementary Note 7. Competing agent details

**OpenAI Advanced Data Analysis:** We accessed this agent through the February 2025 version of ChatGPT Plus (https://chatgpt.com). ChatGPT Plus is a paid subscription service ($20 per month as of February 2025). ChatGPT's user interface supports CSV file upload.

**Open Interpreter:** We used version v0.4.2 of Open Interpreter (https://github.com/OpenInterpreter/open-interpreter). Open Interpreter's system prompt was appended with "Run shell commands with -y so the user doesn't have to confirm them" following the documentation's suggestion. Open Interpreter was set to offline mode.

**Data Interpreter:** We accessed Data Interpreter through version v0.8.1 of MetaGPT (https://github.com/geekan/MetaGPT).

**LAMBDA:** We accessed LAMBDA through commit number 48725f8 of the official GitHub implementation (https://github.com/AMA-CMFAI/LAMBDA). Since the code is designed for the user interface and is incompatible with Python scripting, relative imports were manually added to the cloned LAMBDA repository to enable script-based evaluation.

**Note regarding Data Interpreter:** Based on the Data Interpreter documentation, we were unable to configure the agent to perform conversational data analysis inherently. Specifically, we found that the Data Interpreter could only complete a single task before requiring reinitialization. When we attempted to maintain a persistent Data Interpreter instance for each dataset, subsequent queries after the first one consistently triggered the error: "AttributeError: 'NoneType' object has no attribute 'name'. No answer generated."

To work around this limitation, we reinitialized the Data Interpreter object after each response. However, this approach inherently prevents the interpreter from retaining any memory of past data transformations, limiting its ability to perform contextual analysis across multiple queries.

**Supplementary Note 8. DataAnalysisQA benchmark prompts and templates.**

**ChatDA system prompt:** “You are a helpful assistant. You have access to tools for data analysis. If you need to transform data, use your (non-Python) tools to do so. Use your tools to help the user analyze the dataset.”

**ChatDA (tools-only) system prompt:** “You are a helpful assistant. You have access to tools for data analysis. Use your tools to help the user analyze the dataset.”

**OpenAI Advanced Data Analysis initial instruction:** “I am attaching a dataset. I will then give you instructions for analyzing the dataset. Here are some rules: (1) Immediately split the dataset into 80/20 train/test sets using sklearn’s train_test_split function, with random seed 42. (2) If you are explicitly asked to transform the dataset (e.g., scaling, imputation, feature engineering), keep the changes for future questions. (3) When transforming data (e.g., feature scaling), always fit on the train dataset and transform the test dataset based on the train dataset. (4) When fitting models, always fit on the train dataset and predict on the test dataset. (5) For exploratory analysis (e.g., statistical testing, summary statistics), always consider the entire dataset. (6) Temporarily drop rows with missing values in variables of interest prior to each analysis step. (7) Return a sentence for each query describing your findings, round numeric answers to 3 decimal places. (8) Use significance level 0.05 for statistical tests.”

**Open Interpreter initial instruction:** “I am providing you with a dataset located at path {file_path}.\n I will then give you instructions for analyzing the dataset. Here are some rules: (1) Immediately split the dataset into 80/20 train/test sets using sklearn’s train_test_split function, with random seed 42. (2) If you are explicitly asked to transform the dataset (e.g., scaling, imputation, feature engineering), keep the changes for future questions. (3) When transforming data (e.g., feature scaling), always fit on the train dataset and transform the test dataset based on the train dataset. (4) When fitting models, always fit on the train dataset and predict on the test dataset. (5) For exploratory analysis (e.g., statistical testing, summary statistics), always consider the entire dataset. (6) Temporarily drop rows with missing values in variables of interest prior to each analysis step. (7) Return a sentence for each query describing your findings, round numeric answers to 3 decimal places. (8) Use significance level 0.05 for statistical tests.”

**Data Interpreter task prompt:** “I am providing you with a dataset located at path {file_path}.\n I will then give you instructions for analyzing the dataset. Here are some rules: (1) Immediately split the dataset into 80/20 train/test sets using sklearn’s train_test_split function, with random seed 42. (2) If you are explicitly asked to transform the dataset (e.g., scaling, imputation, feature engineering), keep the changes for future questions. (3) When transforming data (e.g., feature scaling), always fit on the train dataset and transform the test dataset based on the train dataset. (4) When fitting models, always fit on the train dataset and predict on the test dataset. (5) For exploratory analysis (e.g., statistical testing, summary statistics), always consider the entire dataset. (6) Temporarily drop rows with missing values in variables of interest prior to each analysis step. (7) Return a sentence for each query describing your findings, round numeric answers to 3 decimal places. (8) Use significance level 0.05 for statistical tests.\n {task}"

**LAMBDA initial instruction:** "I am providing you with a dataset. I will then give you instructions for analyzing the dataset. Here are some rules: (1) Immediately split the dataset into 80/20 train/test sets using sklearn's train_test_split function, with random seed 42. (2) If you are explicitly asked to transform the dataset (e.g., scaling, imputation, feature engineering), keep the changes for future questions. (3) When transforming data (e.g., feature scaling), always fit on the train dataset and transform the test dataset based on the train dataset. (4) When fitting models, always fit on the train dataset and predict on the test dataset. (5) For exploratory analysis (e.g., statistical testing, summary statistics), always consider the entire dataset. (6) Temporarily drop rows with missing values in variables of interest prior to each analysis step. (7) Return a sentence for each query describing your findings, round numeric answers to 3 decimal places. (8) Use significance level 0.05 for statistical tests."

**Supplementary Note 9. Machine learning benchmark prompts and templates.**

**ChatDA system prompt:** “You are a helpful assistant. You have access to tools for training and evaluating machine learning models. Use your tools to help the user train the best possible model for the given dataset.”

**OpenAI Advanced Data Analysis initial instruction: “**I am attaching a dataset. You must perform a 70/30 train/test split using sklearn’s train_test_split function, with random seed 42. I will then give you instructions for next steps.”

**Open Interpreter task prompt:** “The dataset is at {file_path}.\n You must perform a 70/30 train/test split using sklearn’s train_test_split function, with random seed 42.\n Predict the variable `{target}` with machine learning {task}. Please train the best possible model to accomplish this task. Report the test {metric} of the best possible model you can train. Only report the test {metric} value, rounded to 3 decimal points.”

**Data Interpreter task prompt:** “The dataset is at {file_path}.\n You must perform a 70/30 train/test split using sklearn’s train_test_split function, with random seed 42.\n Predict the variable `{target}` with machine learning {task}. Please train the best possible model to accomplish this task. Report the test {metric} of the best possible model you can train. Only report the test {metric} value, rounded to 3 decimal points.”

**LAMBDA initial instruction:** “You must perform a 70/30 train/test split using sklearn’s train_test_split function, with random seed 42.\n Predict the variable `{target}` with machine learning {task}. Please train the best possible model to accomplish this task. Report the test {metric} of the best possible model you can train. Only report the test {metric} value, rounded to 3 decimal points.”

**Supplementary Note 10. Clinical case study knee arthroplasty dataset details.**

A total of 1,419 patients between the ages of 34 and 92 were included within this study, with full demographics detailed in Supplementary Table 4. A total of 1,419 CT scans of the knee for preoperative Total Knee Arthroplasty (TKA) procedures were provided in Digital Imaging and Communications in Medicine standard format with the same CT protocol from a database across 4 different surgeons at a single multicenter referral center. Quantitative image features (i.e., the angle and width measurements listed in Supplementary Table 4) were computed following the procedures described by Woo et al.[11]. A fellowship-trained orthopedic surgeon labeled whether each knee could have qualified as a Unicompartmental Knee Arthroplasty (UKA) candidate based on differences in compartmental osteoarthritis and morphological changes. The orthopedic surgeon was blinded to the sex, age, and medical history of the patient.

The dataset was de-identified according to the "Safe Harbor" guidelines (https://www.hhs.gov/hipaa/for-professionals/special-topics/de-identification/index.html) prior to being analyzed by ChatDA (GPT-4o, tools-only mode).

**Supplementary Note 11. Clinical case study conversation transcript.**

The data analysis was conducted through the ChatDA user interface. 30% of the data was withheld for model evaluation. For this conversation, ChatDA ran on GPT-4o with code execution disabled (tools-only mode) to minimize the risk of individual data re-identification (see Main for details). Multimodal capabilities were also disabled for this analysis.

**User:** Hi there! Can you provide an overview of the dataset we're working with?

**ChatDA:** Certainly! Here's a quick overview of your dataset:

- Training Set:
  - Number of Rows: 993
  - Number of Variables: 36
  - Categorical Variables: 5
  - Numeric Variables: 31
- Test Set:
  - Number of Rows: 426
  - Number of Variables: 36
  - Categorical Variables: 5
  - Numeric Variables: 31
- Numeric Variables: These include measurements and angles related to the femur, tibia, and other anatomical features, as well as demographic information like age, height, and weight.
- Categorical Variables: These include classifications and characteristics such as arthroplasty classification, external rotation, laterality, sex, and varus alignment.

If you have any specific questions or need further analysis on any part of the dataset, feel free to ask!

**User:** Please list the variable names for me.

**ChatDA:** Here are the variable names in your dataset:

Numeric Variables:

1. AP_Femur_Distance
2. AP_Tibial_Distance
3. Acetabular_Angle
4. Age
5. Femoral_Anteversion
6. Femoral_Head_Width
7. Femoral_Neck_Angle
8. Femoral_Torsion
9. Height
10. Hip-Knee-Ankle_Angle
11. Joint_Line_Convergence_Angle
12. Knee_Flexion_Angle
13. Lateral_Distal_Femoral_Angle
14. Lateral_Femoral_Condylar_Width
15. Lateral_Posterior_Tibial_Slope

16. Lateral_Tibial_Width
17. Medial_Femoral_Condylar_Width
18. Medial_Posterior_Tibial_Slope
19. Medial_Proximal_Tibial_Angle
20. Medial_Tibial_Width
21. Neck-Shaft_Angle
22. Patella_Width
23. Posterior_Condylar_Angle
24. Sulcus_Angle
25. TEA-PCA_Angle
26. TEA_Horizontal_Angle
27. TT-TG_Distance
28. Tibial_Width
29. Tibiofemoral_Angle
30. Trochlear_Groove_Width
31. Weight

Categorical Variables:

1. Arthroplasty_Classification
2. External_Rotation
3. Laterality
4. Sex
5. Varus_Alignment

If you need more details or analysis on any specific variable, just let me know!

**User:** My overall goal is to predict the type of arthroplasty and to understand what variables are predictive of arthroplasty type. What steps should we take to achieve this?

**ChatDA:** To achieve your goal of predicting the type of arthroplasty and understanding the predictive variables, we can follow these steps:

1. Data Preprocessing:
   - Handle Missing Values: Impute or drop missing values as necessary.
   - Scale Numeric Variables: Standardize or normalize numeric variables to ensure they are on a similar scale.
   - Encode Categorical Variables: Convert categorical variables into a format suitable for machine learning models (e.g., one-hot encoding).
2. Exploratory Data Analysis (EDA):
   - Visualize Distributions: Plot distributions of numeric variables and value counts of categorical variables.
   - Correlation Analysis: Check correlations between numeric variables and the target variable to identify potential predictors.
3. Feature Selection:
   - Use feature selection techniques to identify the most important variables for predicting the type of arthroplasty.
4. Model Building:
   - Classification Models: Train various classification models (e.g., Logistic Regression, Random Forest, XGBoost) to predict the type of arthroplasty.

    - Hyperparameter Tuning: Optimize model hyperparameters for better performance.
5. Model Evaluation:
    - Evaluate model performance using metrics such as accuracy, precision, recall, and F1-score.
    - Use cross-validation to ensure the model's robustness.
6. Interpretation:
    - Analyze feature importances from the best-performing model to understand which variables are most predictive of arthroplasty type.
7. Deployment (if applicable):
    - Prepare the model for deployment if you plan to use it in a production environment.

Would you like to start with any specific step, or do you have any particular area you'd like to focus on first?

**User:** That sounds good. Maybe we can start by learning more about the distribution of the arthroplasty type.

**ChatDA:** The distribution of the arthroplasty types in your dataset is as follows:

- TKA (Total Knee Arthroplasty): 471 instances
- UKA (Unicompartmental Knee Arthroplasty): 324 instances

This gives us a good starting point to understand the balance between the two classes. If you have any further questions or would like to proceed with another step, let me know!

**User:** Hmm… how come there are fewer total values than the number of observations in the dataset?

**ChatDA:** It seems there might be some missing values in the Arthroplasty_Classification variable, which could explain why the total count of TKA and UKA is less than the total number of observations in the dataset.

Would you like to address these missing values, or is there anything else you'd like to explore?

**User:** Ah. That makes sense. What percentage of observations is missing in the Arthroplasty classification variable?

**ChatDA:** The Arthroplasty_Classification variable has a missing rate of approximately 43.97%, which means that about 624 observations are missing this classification.

Would you like to address these missing values, or is there another aspect of the dataset you'd like to explore?

**User:** How should we handle the missingness?

**ChatDA:** Handling missing data is crucial for building robust models. Here are a few strategies you can consider for dealing with the missing values in the Arthroplasty_Classification variable:

1. Imputation:

- Predictive Imputation: Use other variables to predict the missing values. This can be done using models like logistic regression or decision trees.
- Simple Imputation: Impute missing values with the most frequent category (TKA in this case), though this might introduce bias.

2. Data Reduction:
   - Remove Missing Observations: If the missing data is not random and constitutes a significant portion of the dataset, you might consider removing these observations. However, this could lead to a loss of valuable data.
3. Modeling with Missing Data:
   - Some machine learning models can handle missing data directly, such as tree-based models (e.g., Random Forests, XGBoost).
4. Analysis of Missingness:
   - Investigate if the missingness is related to other variables, which might provide insights into the nature of the missing data.

Would you like to proceed with any of these strategies, or do you have a specific approach in mind?

**User:** I think imputation isn't the best approach since we are trying to predict the arthroplasty classification variable to begin with. Let's proceed with the data reduction approach.

**ChatDA:** I've removed the observations with missing values in the Arthroplasty_Classification variable. Here's the updated dataset shape:

- Training Set:
  - Before: 993 rows
  - After: 556 rows
- Test Set:
  - Before: 426 rows
  - After: 239 rows

With the missing data addressed, we can now proceed with further analysis or model building. Let me know what you'd like to do next!

**User:** Can you help me visualize the arthroplasty classification variable?

**ChatDA:** The distribution of the Arthroplasty_Classification variable has been plotted. You can view the visualization to see the balance between the TKA and UKA classes. If you have any further questions or need additional analysis, feel free to ask!

**User:** Help me visualize pairwise relationships between Age, Weight, Sex, and Arthroplasty classification.

**ChatDA:** The pairwise relationships between Age, Weight, Sex, and Arthroplasty_Classification have been visualized in a pairplot. You can view the plot to explore these relationships further. If you have any questions or need more insights, just let me know!

**User:** Can you help me figure out if there's a statistically significant relationship between the arthroplasty classification variable and sex?

**ChatDA:** The Chi-squared test was conducted to examine the relationship between Arthroplasty_Classification and Sex. Here are the results:

- Chi-squared Statistic: 2.74
- p-value: 0.098
- Degrees of Freedom: 1

The p-value is greater than the common significance level of 0.05, suggesting that there is no statistically significant relationship between arthroplasty classification and sex. Therefore, we fail to reject the null hypothesis that these two variables are independent.

If you have any further questions or need additional analysis, feel free to ask!

**User:** Cool. Can you list the variables for me again?

**ChatDA:** Certainly! Here are the variable names in your dataset:

Numeric Variables:

1. AP_Femur_Distance
2. AP_Tibial_Distance
3. Acetabular_Angle
4. Age
5. Femoral_Anteversion
6. Femoral_Head_Width
7. Femoral_Neck_Angle
8. Femoral_Torsion
9. Height
10. Hip-Knee-Ankle_Angle
11. Joint_Line_Convergence_Angle
12. Knee_Flexion_Angle
13. Lateral_Distal_Femoral_Angle
14. Lateral_Femoral_Condylar_Width
15. Lateral_Posterior_Tibial_Slope
16. Lateral_Tibial_Width
17. Medial_Femoral_Condylar_Width
18. Medial_Posterior_Tibial_Slope
19. Medial_Proximal_Tibial_Angle
20. Medial_Tibial_Width
21. Neck-Shaft_Angle
22. Patella_Width
23. Posterior_Condylar_Angle
24. Sulcus_Angle
25. TEA-PCA_Angle
26. TEA_Horizontal_Angle
27. TT-TG_Distance
28. Tibial_Width
29. Tibiofemoral_Angle
30. Trochlear_Groove_Width
31. Weight

Categorical Variables:

1. Arthroplasty_Classification
2. External_Rotation
3. Laterality
4. Sex
5. Varus_Alignment

If you need more details or analysis on any specific variable, just let me know!

**User:** I want to proceed with feature selection to predict the arthroplasty classification. Can you help me find the most important features?

**ChatDA:** The feature selection process identified Lateral_Tibial_Width as the most important predictor for the Arthroplasty_Classification variable. You might consider using this feature in your predictive models.

If you'd like to proceed with model building or need further analysis, just let me know!

**User:** Please plot Lateral_Tibial_Width versus Arthroplasty_Classification.

**ChatDA:** The box plot of Lateral_Tibial_Width versus Arthroplasty_Classification has been generated. You can view the plot to explore the relationship between these variables. If you have any further questions or need additional insights, feel free to ask!

**User:** Can you provide me with summary statistics for all the variables?

**ChatDA:** Here are the summary statistics for the variables in your dataset:

Numeric Variables:

- AP_Femur_Distance: Mean = 60.67, Std = 5.30, Missing = 7
- AP_Tibial_Distance: Mean = 50.30, Std = 4.93, Missing = 5
- Acetabular_Angle: Mean = 7.29, Std = 7.27, Missing = 7
- Age: Mean = 67.20, Std = 8.61, Missing = 5
- Femoral_Anteversion: Mean = 11.54, Std = 9.50, Missing = 20
- Femoral_Head_Width: Mean = 44.27, Std = 6.14, Missing = 35
- Femoral_Neck_Angle: Mean = 10.12, Std = 9.14, Missing = 15
- Femoral_Torsion: Mean = 12.65, Std = 7.63, Missing = 24
- Height: Mean = 1.66, Std = 0.10, Missing = 662
- Hip-Knee-Ankle_Angle: Mean = 173.41, Std = 4.25, Missing = 11
- Joint_Line_Convergence_Angle: Mean = 2.68, Std = 1.76, Missing = 11
- Knee_Flexion_Angle: Mean = 7.68, Std = 5.99, Missing = 15
- Lateral_Distal_Femoral_Angle: Mean = 8.87, Std = 3.57, Missing = 9
- Lateral_Femoral_Condylar_Width: Mean = 8.87, Std = 2.87, Missing = 4
- Lateral_Posterior_Tibial_Slope: Mean = 3.84, Std = 3.84, Missing = 5
- Lateral_Tibial_Width: Mean = 29.78, Std = 3.4, Missing = 7
- Medial_Femoral_Condylar_Width: Mean = 3.28, Std = 2.99, Missing = 4
- Medial_Posterior_Tibial_Slope: Mean = 5.07, Std = 4.38, Missing = 5
- Medial_Proximal_Tibial_Angle: Mean = 8.56, Std = 3.84, Missing = 12
- Medial_Tibial_Width: Mean = 29.00, Std = 3.8, Missing = 7
- Neck-Shaft_Angle: Mean = 128.58, Std = 4.43, Missing = 10
- Patella_Width: Mean = 43.95, Std = 4.09, Missing = 6

- Posterior_Condylar_Angle: Mean = 7.39, Std = 5.87, Missing = 5
- Sulcus_Angle: Mean = 135.77, Std = 8.42, Missing = 5
- TEA-PCA_Angle: Mean = 5.07, Std = 2.27, Missing = 5
- TEA_Horizontal_Angle: Mean = 7.68, Std = 6.06, Missing = 5
- TT-TG_Distance: Mean = 12.17, Std = 4.95, Missing = 5
- Tibial_Width: Mean = 76.77, Std = 6.3, Missing = 7
- Tibiofemoral_Angle: Mean = 4.69, Std = 4.21, Missing = 7
- Trochlear_Groove_Width: Mean = 36.40, Std = 4.37, Missing = 5
- Weight: Mean = 85.59, Std = 17.59, Missing = 6

Categorical Variables:

- Arthroplasty_Classification: Most common = TKA, Missing = 0
- External_Rotation: Most common = False, Missing = 16
- Laterality: Most common = right, Missing = 0
- Sex: Most common = F, Missing = 0
- Varus_Alignment: Most common = True, Missing = 8

If you need more detailed insights or further analysis, feel free to ask!

**User:** Can you please impute the missing values in the dataset? Then, please re-perform the feature selection process.

**ChatDA:** To proceed with imputing the missing values, I need to know the strategies you'd like to use for numeric and categorical variables. Here are the options:

- For numeric variables: 'mean', 'median', '5nn', '10nn'
- For categorical variables: 'most_frequent', 'missing'

Once you provide these details, I can proceed with the imputation and then re-perform the feature selection process.

**User:** What do these strategies mean? What would you recommend?

**ChatDA:** Here's a brief explanation of each strategy:

Numeric Variables:

1. Mean: Replaces missing values with the mean of the column.
2. Median: Replaces missing values with the median of the column.
3. 5nn: Uses the average of the 5 nearest neighbors to impute missing values.
4. 10nn: Uses the average of the 10 nearest neighbors to impute missing values.

Categorical Variables:

1. Most Frequent: Replaces missing values with the most frequent category in the column.
2. Missing: Treats missing values as a separate category.

Recommendations:

- For numeric variables, if the data is normally distributed, using the mean is a common choice. If the data has outliers, the median might be more robust.
- For categorical variables, using the most frequent category is generally a good choice unless you want to specifically track missing data as a separate category.

Would you like to proceed with these recommendations, or do you have specific preferences?

**User:** Yes, let's proceed with your recommendations.

**ChatDA:** The missing values in the dataset have been successfully imputed using the mean for numeric variables and the most frequent category for categorical variables. Now, let's proceed with the feature selection process. Could you please specify the target variable and the predictors you want to consider for feature selection?

**User:** Sounds great. We're predicting the Arthroplasty classification variable, as a reminder. Please consider all the other variables in the dataset as potential predictors.

**ChatDA:** The feature selection process identified the following variables as the most important predictors for the Arthroplasty_Classification:

- Joint_Line_Convergence_Angle
- Lateral_Tibial_Width
- Medial_Proximal_Tibial_Angle
- TT-TG_Distance
- Tibial_Width
- Tibiofemoral_Angle

You can consider using these features for your classification model to predict Arthroplasty_Classification. If you need further analysis or model building, feel free to ask!

**User:** I'd like to predict the arthroplasty classification using these predictor variables. Please start with a simple linear model. I want you to interpret the coefficients for me.

**ChatDA:** The logistic regression model has been fitted to predict the Arthroplasty_Classification using the selected predictors. Here's a summary of the model's coefficients and their interpretations:

Coefficients:

- Joint_Line_Convergence_Angle:
    - Estimate: 0.134
    - p-value: 0.009
    - Interpretation: A higher Joint Line Convergence Angle is associated with a higher likelihood of the positive class (UKA). The effect is statistically significant.
- Lateral_Tibial_Width:
    - Estimate: -0.015

    - p-value: 0.708
    - Interpretation: Lateral Tibial Width has a negative but not statistically significant effect on the likelihood of the positive class.
- Medial_Proximal_Tibial_Angle:
    - Estimate: -0.012
    - p-value: 0.633
    - Interpretation: Medial Proximal Tibial Angle has a negative but not statistically significant effect on the likelihood of the positive class.
- TT-TG_Distance:
    - Estimate: -0.041
    - p-value: 0.026
    - Interpretation: A higher TT-TG Distance is associated with a lower likelihood of the positive class. This effect is statistically significant.
- Tibial_Width:
    - Estimate: -0.008
    - p-value: 0.710
    - Interpretation: Tibial Width has a negative but not statistically significant effect on the likelihood of the positive class.
- Tibiofemoral_Angle:
    - Estimate: -0.085
    - p-value: 0.002
    - Interpretation: A higher Tibiofemoral Angle is associated with a lower likelihood of the positive class. This effect is statistically significant.

Model Performance:

- Training Accuracy: 59.7%
- Test Accuracy: 60.7%
- ROC AUC (Test): 0.632

The model shows moderate performance, with some predictors having significant effects. If you want to explore further or try different models, let me know!

**User:** How can I interpret the joint line convergence angle coefficient? What happens when that angle increases by 1?

**ChatDA:** In logistic regression, the coefficients represent the change in the log-odds of the outcome for a one-unit increase in the predictor variable, holding all other variables constant.

For the Joint_Line_Convergence_Angle:

- Coefficient: 0.134

Interpretation:

- A one-unit increase in the Joint Line Convergence Angle increases the log-odds of the outcome being the positive class (UKA) by 0.134, assuming all other variables remain constant.

Odds Ratio:

- To interpret this in terms of odds, you can exponentiate the coefficient:

- Odds Ratio: ($e^{0.134} \approx 1.143$)

This means that for each one-unit increase in the Joint Line Convergence Angle, the odds of the outcome being UKA increase by approximately 14.3%, holding all other factors constant.

This positive relationship indicates that as the Joint Line Convergence Angle increases, the likelihood of the classification being UKA also increases.

**User:** Can you train machine learning models to predict the arthroplasty classification with the same six predictor variables?

**ChatDA:** The machine learning models have been trained to predict Arthroplasty_Classification using the selected predictors. Here's a summary of their performances:

Model Performances:

- Logistic Regression:
  - Training Accuracy: 59.9%
  - Test Accuracy: 60.3%
  - Test ROC AUC: 0.632
- Random Forest (RF):
  - Training Accuracy: 92.3%
  - Test Accuracy: 55.6%
  - Test ROC AUC: 0.584
- XGBoost:
  - Training Accuracy: 87.2%
  - Test Accuracy: 56.9%
  - Test ROC AUC: 0.592

Feature Importances:

- Random Forest: Tibiofemoral_Angle (18.1%), Joint_Line_Convergence_Angle (17.0%), TT-TG_Distance (16.9%)
- XGBoost: Tibiofemoral_Angle (20.7%), Tibial_Width (16.9%), TT-TG_Distance (16.0%)

Interpretation:

- The logistic regression model provides a baseline with moderate performance.
- Both Random Forest and XGBoost show high training accuracy but lower test accuracy, indicating potential overfitting.
- Tibiofemoral_Angle appears to be a significant predictor across models.

If you want to further refine these models or explore other techniques, feel free to ask!

**Supplementary Note 12. Agent software usability.**

We assess agent software usability through five criteria:

1. User interface availability through software
2. Open-source software
3. Accessibility of clear analysis steps and generated code
4. Availability of command line/code-based scripting analysis options
5. Flexibility of analyses

ChatDA received a score of 4 as its analyses are largely limited to its predefined tools (Supplementary Note 2), but all other criteria are satisfied.

OpenAI Advanced Data Analysis received a score of 4 as it is not open source, meaning that users are subject to OpenAI's cloud rate limits, but all other criteria are satisfied.

Data Interpreter received a score of 3 as it can perform flexible analyses, is open source, and has a command line/code-based scripting option. Data Interpreter does not provide a user interface or easy access to analysis steps and generated code.

Open Interpreter received a score of 4 as it lacks a user interface, but all other criteria are satisfied.

LAMBDA received a score of 4 as it does not offer a command line/code-based scripting option, but all other criteria are satisfied.

#### References


1. Grinsztajn, L., Oyallon, E. & Varoquaux, G. Why do tree-based models still outperform deep learning on typical tabular data? in *Proceedings of the 36th International Conference on Neural Information Processing Systems* 507–520 (Curran Associates Inc., Red Hook, NY, USA, 2022).
2. Cheng, A. C. *et al.* Supporting rapid innovation in research data capture and management: the REDCap external module framework. *J. Am. Med. Inform. Assoc.* **32**, 1149–1156 (2025).
3. Narayan, S. M., Kohli, N. & Martin, M. M. Addressing contemporary threats in anonymised healthcare data using privacy engineering. *NPJ Digit. Med.* **8**, 145 (2025).
4. Mao, Y., Stevanoski, B. & Montjoye, Y.-A. de. DeSIA: Attribute Inference Attacks Against Limited Fixed Aggregate Statistics. Preprint at https://doi.org/10.48550/arXiv.2504.18497 (2025).
5. Rocher, L., Hendrickx, J. M. & de Montjoye, Y.-A. Estimating the success of re-identifications in incomplete datasets using generative models. *Nat. Commun.* **10**, 3069 (2019).
6. Naveed, M., Kamara, S. & Wright, C. V. Inference Attacks on Property-Preserving Encrypted Databases. in *Proceedings of the 22nd ACM SIGSAC Conference on Computer and Communications Security* 644–655 (Association for Computing Machinery, New York, NY, USA, 2015). doi:10.1145/2810103.2813651.
7. Escobar, F. A., Canard, S., Laguillaumie, F. & Phan, D. H. Computational Differential Privacy for Encrypted Databases Supporting Linear Queries. *Proc. Priv. Enhancing Technol.* (2024).

8. Sun, M. *et al.* LAMBDA: A Large Model Based Data Agent. *J. Am. Stat. Assoc.* **0**, 1–13.

9. Hu, X. *et al.* InfiAgent-DABench: Evaluating Agents on Data Analysis Tasks. in *Proceedings of the 41st International Conference on Machine Learning* 19544–19572 (PMLR, 2024).

10. Hong, S. *et al.* Data Interpreter: An LLM Agent For Data Science. Preprint at https://doi.org/10.48550/arXiv.2402.18679 (2024).

11. Woo, J. J. *et al.* Who Are the Anatomic Outliers Undergoing Total Knee Arthroplasty? A Computed Tomography–Based Analysis of the Hip-Knee-Ankle Axis Across 1,352 Preoperative Computed Tomographies Using a Deep Learning and Computer Vision–Based Pipeline. *J. Arthroplasty* **39**, S188–S199 (2024).
